# Timeliness of provisional United States mortality data releases during the COVID-19 pandemic: delays associated with electronic death registration system and weekly mortality

**DOI:** 10.1101/2021.01.07.21249401

**Authors:** Janet E. Rosenbaum, Marco Stillo, Nathaniel Graves, Roberto Rivera

## Abstract

All-cause mortality counts allow public health authorities to identify populations experiencing excess deaths from pandemics, natural disasters, and other emergencies. Delays in the completeness of mortality counts may contribute to misinformation because death counts take weeks to become accurate. We estimate the timeliness of all-cause mortality releases during the Covid-19 pandemic for the dates 3 April to 5 September 2020 by estimating the number of weekly data releases of the NCHS Fluview Mortality Surveillance System until mortality comes within 99% of the counts in the 19 March 19 2021 provisional mortality data release. States’ mortality counts take 5 weeks at median (interquartile range 4--7 weeks) to completion. The fastest states were Maine, New Hampshire, Vermont, New York, Utah, Idaho, and Hawaii. States that had not adopted the electronic death registration system (EDRS) were 4.8 weeks slower to achieve complete mortality counts, and each weekly death per 10^8 was associated with a 0.8 week delay. Emergency planning should improve the timeliness of mortality data by improving state vital statistics digital infrastructure.

## Introduction

Mortality is often underestimated for pandemics, natural disasters, and other emergencies (1), but estimated excess mortality can yield a more complete assessment of the mortality impact. Excess mortality can be estimated using statistical models to evaluate whether the number of deaths during the pandemic is greater than would be expected from past mortality patterns by comparing the actual number of deaths for each week (or any other increment) with the number of expected deaths for each week, based on the population, time of year, and secular mortality trends. If excess mortality estimates exceed the official death count from the pandemic, that suggests that the official death count is an under-estimate. Excess mortality greater than the official death counts has been observed from causes including influenza (2), extreme temperatures (3), and hurricanes (4). During pandemics, natural disasters, and other emergencies, policymakers can use estimates of excess mortality to identify populations at greatest risk. Accurate and timely estimation of excess mortality allows policymakers and clinicians to formulate appropriate policy and clinical responses quickly enough for these responses to save lives.

During the Covid-19 pandemic, the U.S. Centers for Disease Control and Prevention found that excess mortality exceeded the official Covid-19 mortality count (5). Covid-19 deaths may have been under-counted for several reasons, including underdiagnosed Covid-19 due to low test access, lack of surveillance testing (6), atypical disease presentation, sudden Covid-19 declines (7), sudden Covid-19 deaths (8), not seeking care because many Covid-19 patients did not perceive hypoxia and lung damage (9), or etiologically nonspecific death reporting (10) due to guidelines that limited post-mortem testing (11). Additional excess deaths may have been due to delays in seeking healthcare for acute non-Covid-19 conditions, such as stroke or heart attack (12). Under-counting deaths permitted the minimization of the extent of the Covid-19 pandemic (13). Timely and accurate excess death estimates could be important tools to combat disinformation (13), encourage non-pharmaceutical interventions (14), and also inform about the importance of seeking health care, for both acute non-Covid-19 illnesses and for Covid-19.

In the United States, public health statisticians often estimate excess mortality from weekly provisional all-cause mortality data from the National Vital Statistics System, that exclude deaths not yet reported and are updated in successive weekly releases (15). States differ from each other in the timeliness of death reporting, in part because states vary in the extent of adoption of the Electronic Death Registration System (16). Timeliness of death reporting has improved in recent years: within 13 weeks, all-cause death data were 84% complete in 2015 (15) and 95% complete in 2017 (17). We estimated the time until all-cause mortality counts for each state are complete. Past research does not explore a variety of reasons for the timeliness of provisional mortality estimates, so in an exploratory analysis, we also evaluated potential explanations for timeliness, such as the extent of electronic death registration adoption, death investigation system, weekly mortality, and state resources measured by GDP and public health budget. Public health authorities and researchers can rapidly estimate excess mortality for a variety of emergencies using weekly all-cause mortality, which makes this measure important.

## Methods

### Data

We archived 35 weeks of provisional mortality counts by state from the National Vital Statistics System between 3 April and 4 December 2020 distributed by the National Center for Health Statistics Mortality Surveillance System using the Fluview web interface (https://gis.cdc.gov/grasp/fluview/mortality.html). The federal government updates provisional mortality data every Friday on the Fluview web interface. The provisional counts are stratified into 52 jurisdictions: all 50 states and the District of Columbia, with New York City (NYC) and non-NYC New York State separated.

## Measures

### Primary outcome

Our primary outcome is the weeks of delay until mortality counts are complete for each of the 23 weeks from 3 April - 5September 2020. This estimation resulted in 1196 mortality delay observations from 52 jurisdictions. We measure delay as the number of weekly data releases until mortality counts reached at least 99% of the counts in the most recent provisional data release: 19 March 2021. We chose the ending date, 5 September 2020, 12 weeks before the most recent data release at the time of the first analysis, 4 December 2020.

For example, the 17 April 17 2020 release is the first provisional data release for deaths during the week of 3 April 2020. A 4-week delay until completeness would mean that the provisional count of deaths for 3 April exceeded 99% of the count in the most recent provisional release 4 weeks later, on 8 May 2020. We assessed the face validity of these mortality reporting delay estimates by comparison with a spaghetti plot for each jurisdiction, where each line represents a weekly release (Figures S1, S2, S4).

### Primary predictor

The primary predictor variable was adoption of the electronic death registration system (EDRS) prior to the starting point of this data. We assessed EDRS in two forms: as a binary variable and as an ordered categorical variable. In 2020, prior to the pandemic, 4 states did not use electronic death registration (CT, NC, RI, WV); the binary indicator of non-adoption of electronic death registration was coded as 1 for these four states and otherwise 0. In addition to this binary indicator of adoption of electronic death registration, we used an ordered categorical variable from the most recent report of the extent of electronic record adoption assessed in 2018 (16): 9 states have fewer than 75% death certificates filed with electronic death registration (AR, CO, MD, MI, MS, NY, PA, TN, VA), and the 38 remaining jurisdictions (37 states, NYC, and DC) file more than 75% of death certificates with electronic death registration (16). This report did not provide the numerical percentage of death certificates filed electronically in 2018, only these categories. It is reasonable to believe that the closer jurisdictions get to 100% of death certificates filed electronically, the smaller the delay to mortality count completeness. We confirmed electronic death registration implementation with each state’s public health vital statistics website.

### Additional predictor variables

We hypothesized that during weeks with more all-cause deaths, the completeness of mortality counts would have greater delays, due to the resources needed for processing additional deaths; we tested whether weekly deaths or weekly deaths per hundred million were associated with delay. Weekly deaths per population to hundred million ranged from 1-10, with a median of 2, so coefficients were most interpretable on this scale.

We hypothesized that states with more economic resources would have faster death certificate processing because they have more money to upgrade state vital statistics infrastructure. We measured economic resources for the 50 states and the District of Columbia using the Bureau of Economic Analysis’s 2018 per capita GDP; although New York State’s delay excludes NYC death certificates, the tax base of New York State includes NYC. We retrieved the public health budget per million residents from public records and used it as a separate measure of economic resources.

To assess whether our delay measure is associated with a prior measure of data completeness, we used a 2017 measure of the percent of death certificates available within 13 weeks as a covariate (17).

Deaths that occur outside a physician’s supervision require a death investigation process to identify the cause of death (disease or injury and any underlying causes) and whether the death was natural, accidental, homicide, suicide, or undetermined (manner of death). We hypothesized that the death investigation system may be associated with delay. Death investigations may be conducted by medical examiners, who are physicians, or by coroners, who are usually non-physicians with no special qualifications, except in 4 states (Kansas, Louisiana, Minnesota, and Ohio) that require coroners to be physicians. Having a medical examiner or physician coroner is a marker of a professionalized death investigation system (18), so we hypothesized that death investigations conducted by physicians may have fewer delays. States also differ in whether they have centralized offices for death investigations at the state level or decentralized ones at the county or district level. We defined a variable based on the CDC’s coding of death investigation system type (19): centralized (state-level) medical examiner system, county- or district-based medical examiner system, county-based system with a mixture of coroner and medical examiner office, or a county-, district-, or parish-based coroner system. New York City created the first centralized medical examiners system in 1918, so NYC was coded as having county/district medical examiners and having a medical examiner system (18).

We used date as a continuous variable because there may be changes over time. We also evaluated whether the month of the year was associated with delay because states may differ in reporting practices over time, such as if they learned from other states’ experiences. We created binary indicators for the month: April, May, June, July, and August and the first two weeks of September.

### Statistical Analysis

We used Poisson regression with weeks of delay as the outcome variable, with varying intercept by states (20). We plotted these varying intercepts for the null model (Figure 1) (21). The residuals were not overdispersed, based on the estimated dispersion factor for the general linear mixed model (22). We estimated the delay associated with paper-based systems using fixed slope and varying intercept regression models (20). The model used a categorical variable for no adoption, less than 75% adoption, and more than 75% adoption assessed in 2017 (16).

**Figure 1:**
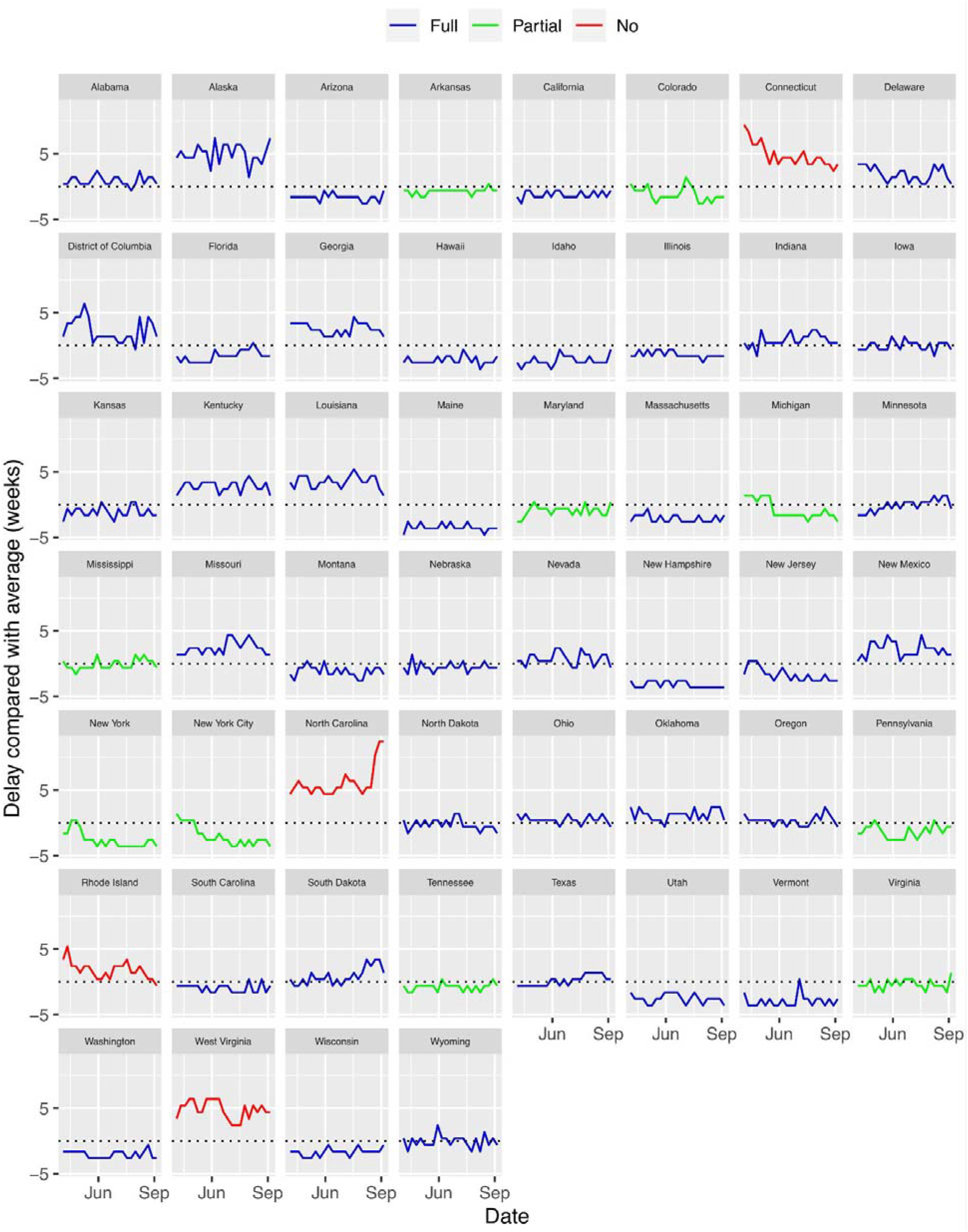
Delay in mortality reporting by date and adoption of electronic death registration system, compared with average (dotted line)

As a robustness check, we repeated the model using only a binary indicator for no electronic death registration adoption, which yielded similar estimates. In exploratory analysis using a log-likelihood ratio test to identify variables that improve the fit of the model, we evaluated additional covariates: weekly mortality per hundred million population, state gross domestic product (GDP) per capita, population, state public health budget per capita, the 4-level death investigation variable, whether the state has a medical examiner, whether a state uses only coroners, the 2017 electronic record submission measure, date, and month of the year. Weekly mortality per hundred million population was associated with delay, but the other variables were not.

This study is an analysis of publicly available data from United States federal sources in broad categories such that individuals cannot be identified, so it is not human subjects research and is exempt from requiring human subjects board review. We have made the raw data and code publicly available through a Github repository: https://github.com/Misreporting/mortality-reporting

All analyses were performed in R 4.0.3 between April and December 2020 with revisions in March 2021.

## Results

On average, all-cause mortality counts take 5.6 weeks to become complete with less than 1% increases subsequently. Figure 1 shows a plot of delay in reporting all-cause mortality count completeness from all 52 jurisdictions, the outcome variable for the regression. Figure 2 shows the average number of weeks of delay until mortality count completeness for all 52 jurisdictions. The slowest states are North Carolina, Alaska, Connecticut, and West Virginia, which are respectively delayed by 12.4, 11.1, 10.9, and 10.9 weeks on average, and the fastest states are Maine, New Hampshire, and Vermont, which are delayed by 2.5, 2.8, and 3.0 weeks, a gap of almost 10 weeks between the slowest and fastest states.

**Figure 2:**
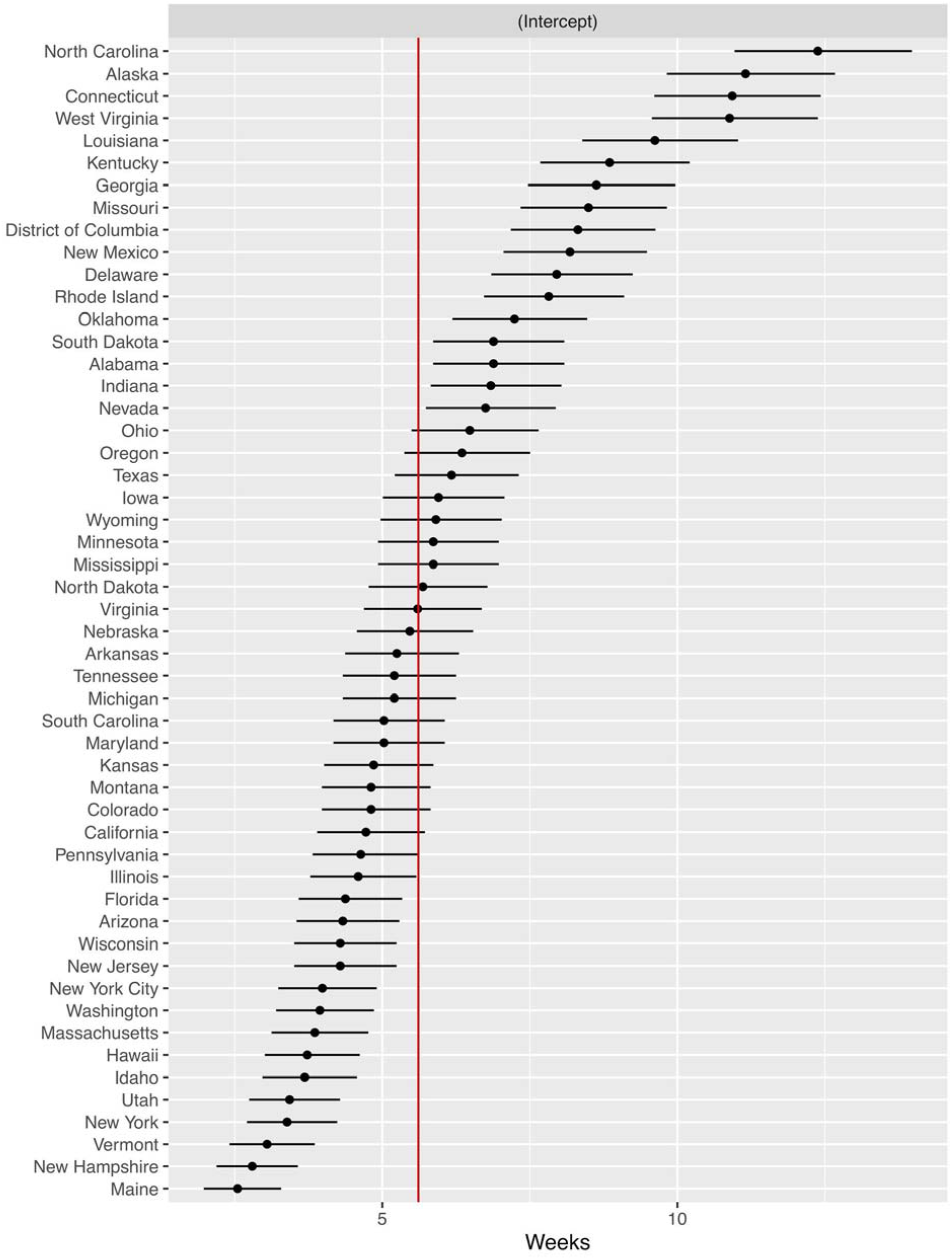
Weeks until all-cause mortality counts are complete for April 3-September 5, 2020. The red line shows the mean delay.

The jurisdictions with quicker than average time until mortality counts are complete were Pennsylvania, Illinois, Florida, Arizona, Wisconsin, New Jersey, New York City (NYC), Washington, Massachusetts, Hawaii, Idaho, Utah, New York State (excluding NYC), Vermont, New Hampshire, and Maine (Figure 2). The states with average time until completeness are Ohio, Oregon, Texas, Iowa, Wyoming, Minnesota, Mississippi, North Dakota, Virginia, Nebraska, Arkansas, Tennessee, Michigan, South Carolina, Maryland, Kansas, Montana, Colorado, and California (Figure 2).

Adjusted for weekly deaths, the jurisdictions that were quicker than average and average were the same as unadjusted for weekly deaths, but the order changed (Figure S3). Jurisdictions with high incidence of Covid-19, such as NYC, had a slightly lower delay adjusted for Covid-19 cases.

Table 1 shows the regression results predicting delay in mortality count completeness with varying intercept by state. Compared with full electronic death registration adoption (greater than 75% of death certificates reported electronically) and controlling for weekly deaths, states without electronic death registration adoption took 85% longer (1.85, 95% confidence interval (1.31, 2.61)), which translates to 4.8 weeks longer. The delay for states with partial electronic death registration did not differ from states with full electronic death registration adoption.

**Table 1:** Poisson regression to predict delay in mortality reporting with varying intercept by state (n=1196 observations of 52 jurisdictions)

|  | IRR | 95% CI | p |
| --- | --- | --- | --- |
| Intercept | 3.96 | (3.42, 4.56) | <0.001 |
| Weekly deaths per 10<sup>8</sup> | 1.14 | (1.09, 1.20) | <0.001 |
| Electronic death registration system |  |  |  |
| Full adoption | Ref. |  |  |
| Partial adoption | 0.84 | (0.66, 1.06) | 0.1 |
| No adoption | 1.85 | (1.31, 2.61) | <0.001 |

IRR = incidence rate ratio, exponentiated coefficients of Poisson regression
95% CI = 95 percent confidence interval

Weekly deaths per 100 million population ranged from 0.9--9.6 with a median of 1.9 deaths per 100 million; the interquartile range was 1.7 to 2.2 weekly deaths per 100 million population. Each additional weekly death per 100 million population was associated with 14% more weeks of delay (95% CI (1.09, 1.20)), which translates to 0.8 more weeks.

All states that did not yet implement EDRS used a centralized state-based medical examiner. Delay is associated with death investigation system type: centralized state medical examiner offices (median (M) 6 weeks, interquartile range (IQR) 3--9 weeks), county-based mixture of medical examiner and coroner offices (M 5 weeks, IQR 4--6 weeks), county/district-based coroner offices (M 5 weeks, IQR 4--6 weeks), and county/district-based medical examiner offices (M 4 weeks, IQR 4--5 weeks) (Kruskal-Wallis test p<0.001) (Figure 3). The association between death investigation system and delay remained after excluding states that did not implement EDRS (Kruskal-Wallis test p<0.001), but there was no association in the Poisson regression with varying intercept by state.

**Figure 3:**
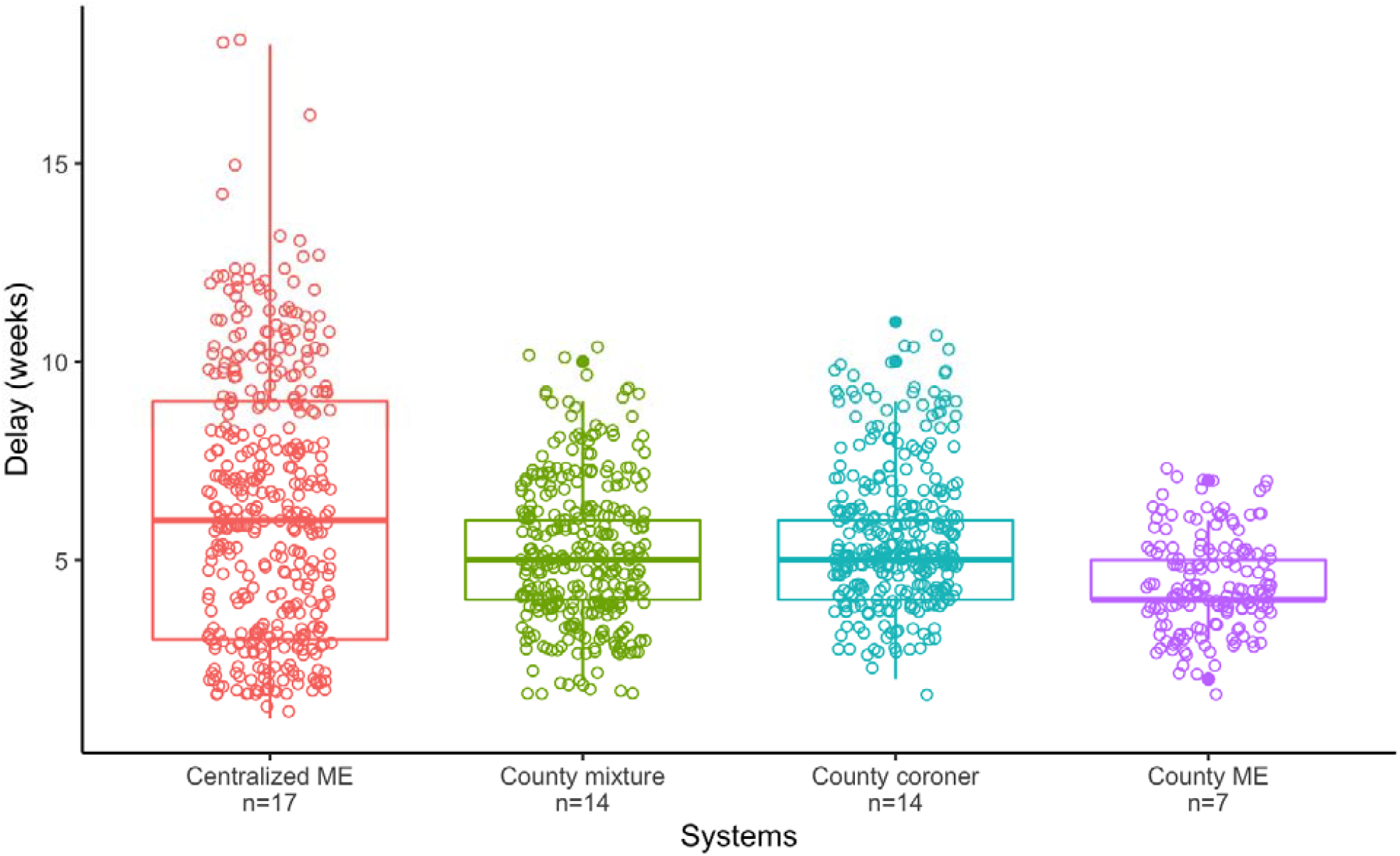
Association between death reporting system and weeks of delay (n=52: 50 states, NYC, and DC). Centralized ME: Centralized state medical examiner office (median (M) 6 weeks, interquartile range (IQR) 3--9 weeks, n=17 states) County mixture: County-based mixture of medical examiner and coroner offices (M 5 weeks, IQR 4--6 weeks, n=14 states) County coroner: County/district-based coroner offices (M 5 weeks, IQR 4--6 weeks, n=14 states) County ME: County/district-based medical examiner offices (M 4 weeks, IQR 4--5 weeks, n=7 states/jurisdictions)

In Poisson regression with varying intercept by state, delay was also not associated with state resources, per capita GDP, per capita public health budget, all-cause mortality completeness within 13 weeks in 2017, population, date, or month, based on likelihood ratio tests of nested models that included these variables.

We performed three robustness checks for our model that did not change the model results substantially. To address slight autocorrelation in the residuals (Durbin-Watson statistic = 1.5), we repeated the analysis using penalized quasi-likelihood with autocorrelation-moving average residuals with a correlation structure of order (p=2, q=2) (23). We identified the autoregression parameter of 2 using the marginally significant lags (0.25 and 0.1) in the partial autocorrelation function plot and the moving average parameter of 2 using the marginally significant lags (0.25 and 0.18) in the autocorrelation function plot. The result changes were negligible: states without electronic death records took 92% longer (1.92, 95% CI (1.37, 2.69)), which translates to 5.2 weeks longer (Table S1). Our measure of EDRS adoption dates to 2018, so as a second robustness check we repeated the analysis with a binary variable for EDRS adoption that was accurate at the time of the data as a robustness check. The result changes were negligible: controlling for weekly deaths, states without electronic death registration took 92% longer (1.92, 95% CI (1.31, 2.64)), which translates to 5.2 weeks longer (Table S2). As a third robustness check, we repeated the analysis using log deaths per log million population and the results changed negligibly. States without electronic death records took 93% longer (1.93, 95% CI (1.33, 2.79)), which translates to 5.2 weeks longer and the number of deaths remained associated with delay (Table S3).

## Discussion

All-cause mortality is a vital public health tool for understanding the true mortality burden of natural disasters and health emergencies, when causes of death may not be coded accurately. Quickly reporting all-cause mortality data can improve public health policy by identifying populations with death burdens larger than the official count in time to intervene with these populations. The large differences in delays between states suggest that many states could improve the timeliness of their all-cause mortality data. Delays in reporting mortality result in provisional counts lower than actual mortality. Perceived risk of disease is an important determinant of health behavior (24), so delays in reaching complete provisional mortality counts may contribute to the pandemic misinformation that Covid-19 mortality was exaggerated (25) and reduce public adherence to non-pharmaceutical interventions such as mask-wearing (26) (14).

These delays in reaching complete mortality counts are not attributable to state resources: high-resource states are no faster than low-resource states. The three slowest states, North Carolina, Connecticut, and Alaska, are the 33rd, 4th, and 8th richest states, and the three fastest states, Maine, Vermont, and New Hampshire, are the 43rd, 36th, and 18th richest states. County-based medical examiner death investigation systems are fastest on average, which may be because medical examiner systems are more professionalized than coroner-based systems (18). State-based medical examiner offices are the slowest at median, so they may be under-staffed relative to county-based offices or require more steps for investigation.

Connecticut and North Carolina began to pilot electronic systems respectively in July 2020 (27) and October 2020 (28). However, our results suggest that substantial delays in all-cause death counts occur even in states that fully implemented electronic death registration. Further, Connecticut’s delays decreased in mid-May when mortality decreased, rather than in July when the electronic system began implementation; among Connecticut’s 5 weeks with the largest delays (12+ weeks), 4 weeks were also the highest mortality weeks.

### Strengths and Limitations

In contrast with the completeness measure disseminated through FluView, this analysis uses a measure of data timeliness that can identify delays in reporting deaths during a period of high mortality. The completeness measure disseminated through FluView compares the number of reported deaths with the average number of deaths from prior years, so the FluView completeness measure is accurate only during periods of average mortality. Although we could not measure the delay in reporting each death --- that is, the time between a death occurred and the death certificate was counted --- we were able to assess the delay until mortality counts came within 1% of the count in the March 19, 2021 provisional mortality release.

It is possible that the delays estimated in this study were due specifically to the Covid-19 pandemic. We do not have access to states’ internal documentation regarding death reporting procedures and we do not know whether states required additional review steps because of the Covid-19 pandemic. After the Covid-19 pandemic, future research can evaluate mortality reporting delays in order to evaluate the need for state reforms to improve timeliness.

Delays are not due only to state-level differences in coding causes of death and reporting these to the NCHS. Delays may be due to differences that occur at the federal level when the National Center for Health Statistics reviews death certificates, ICD-10 codes, and processes data that are reported to the public. NCHS must manually assign ICD-10 codes for new causes of deaths and multiple causes of death, both likely in the case of Covid-19. States with more Covid-19 deaths would be expected to have more federal delays. If these federal delays differ systematically between states, these federal differences could explain the observed delays, not the states themselves (29).

The ordered categorical variable for the extent of adoption of electronic death records dated from 2018, which may explain why states categorized in 2018 as filing less than 75% of death certificates electronically did not differ in mortality count timeliness. However, we verified the binary indicator of non-implementation of electronic death records to be accurate as of the time of the data in 2020, and the results were the same using this variable. Alaska is considered to be a full adopter of electronic death registration (15) with 95% completeness within 13 weeks in 2017 (16), but Alaska was among the slowest states by our measure of number of weeks of delay. Alaska is likely *sui generis* because it is uniquely disadvantaged among US states by the lack of roads to the most remote locations in the state, which may explain the lack of timeliness.

### Public health implications

As suggested after earlier pandemics (30), increasing resources to improve the timeliness of mortality data is necessary for pandemic planning. Improving mortality data timeliness will also benefit natural disaster planning, when excess deaths can be used for mortality estimation. The vital statistics infrastructure is under-funded (31). State and federal pandemic planning should seek resolution for delays in mortality reporting so that all-cause deaths can be used to estimate excess deaths to identify areas and populations in need of additional intervention.

The specific features that make a vital statistics system highly efficient likely include many details we could not measure. Likely, there are many details known primarily to the career civil servants that run state vital statistics systems. States could likely benefit from consulting more efficient but otherwise similar states. For example, Utah has substantially lower delay than 4 of its 6 neighboring states. Funeral directors, who enter demographic information on death certificates, adopted electronic death registration quickly, but medical examiners have lagged (16). California and Arizona allowed electronic death registration submissions by fax machine (16), and our analysis found that these states were faster than average. States that consider unconventional approaches for electronic death registration submission that meet the needs of all stakeholders may have similar success.

The CDC includes percent completeness metrics in the Mortality Surveillance System, defined as the number of deaths divided by the average number of deaths from the most recent 4 years. This completeness measure cannot measure completeness accurately during a period of excess deaths, which is when these measures are most crucial and subject to the most public scrutiny. Data completeness measures that can remain accurate during periods of high mortality may reduce misinformation, such as claims that mortality counts are exaggerated.

All countries can estimate delays in mortality completeness. We estimated the timeliness of mortality data using United States data because delays were noticeable in our analysis of excess mortality during the early Covid-19 pandemic (14). On a global scale, the World Health Organization (WHO) (32), the European monitoring of excess mortality for public health action network (33), data journalists at the *Economist* (34) and *Financial Times* (35), and researchers have estimated country-level excess mortality using all-cause mortality data. The WHO has also estimated mortality data completeness and other markers of adequate vital statistics systems (32). Any entity that estimates excess mortality can use our method to estimate the timeliness of mortality data to identify jurisdictions with large delays in mortality reporting. Our findings suggest even high-GDP jurisdictions may have large delays in mortality reporting and lower-GDP jurisdictions may have timely mortality reporting. In the United States context, the adoption of electronic death registration systems predicted more timely mortality reporting, but the most important factors in other jurisdictions may differ.

## Data Availability

All data produced are available online at https://github.com/Misreporting/mortality-reporting

## Conclusions

This exploratory analysis found that the time for states’ provisional mortality counts to become complete varies greatly between states: the quickest states had complete provisional mortality counts within 4 weeks, and the slowest states took 3 times as long as the fastest states. Three of the slowest states have adopted the electronic death registration systems since collection of these data. Given the importance of provisional mortality counts to understand excess mortality during health emergencies, all states should improve the timeliness of vital statistics reporting by replicating more efficient states with similar characteristics. Funding to improve vital statistics infrastructure should be included in emergency planning budgets because vital statistics systems are crucial for understanding all emergencies that increase mortality.

## Notes

**Conflict of interest statement:** On behalf of all authors, the corresponding author states that there is no conflict of interest.

### Competing Interest Statement

The authors have declared no competing interest.

### Funding Statement

No external funding was received to support this work.

### Author Declarations

This study is an analysis of publicly available data from United States federal sources in broad categories such that individuals cannot be identified, so it is not human subjects research and is exempt from requiring human subjects board review.

### Summary of Updates

Revised according to reviewers' suggestions.

