## Supplementary Information for "Timeliness of provisional United States mortality data releases during the COVID-19 pandemic: delays associated with electronic death registration system and weekly mortality"

Table S1: Poisson regression to predict delay in mortality reporting with varying intercept by state (n=1196 observations of 52 jurisdictions) using correlation structure of order (p=2, q=2).

| | IRR | 95% CI | p |
|---|---|---|---|
| Intercept | 4.14 | (3.67, 4.66) | <0.001 |
| Weekly deaths per 10^8 | 1.11 | (1.08, 1.13) | <0.001 |
| Electronic death registration system | | | |
| Full adoption | Ref. | | |
| Partial adoption | 0.90 | (0.70, 1.14) | 0.4 |
| No adoption | 1.92 | (1.36, 2.69) | <0.001 |

IRR = incidence rate ratio, exponentiated coefficients of Poisson regression
95% CI = 95 percent confidence interval

Table S2: Poisson regression to predict delay in mortality reporting with varying intercept by state (n=1196 observations of 52 jurisdictions) .

| | IRR | 95% CI | p |
|---|---|---|---|
| Intercept | 3.82 | (3.33, 4.39) | <0.001 |
| Weekly deaths per 10^8 | 1.14 | (1.08, 1.20) | <0.001 |
| Electronic death registration system | | | |
| Any adoption (Full or partial) | Ref. | | |
| No adoption | 1.86 | (1.31, 2.64) | <0.001 |

IRR = incidence rate ratio, exponentiated coefficients of Poisson regression
95% CI = 95 percent confidence interval

Table S3: Poisson regression to predict delay in mortality reporting with varying intercept by state (n=1196 observations of 52 jurisdictions) using log deaths per log million population.

| | IRR | 95% CI | p |
|---|---|---|---|
| Intercept | 1.34 | (0.62, 1.07) | 0.5 |
| Log weekly deaths per log million population | 22.4 | (4.02, 136.0) | <0.001 |
| Electronic death registration system | | | |
| Full adoption | Ref. | | |
| Partial adoption | 0.90 | (0.62, 1.07) | 0.1 |
| No adoption | 1.92 | (1.33, 2.79) | <0.001 |

IRR = incidence rate ratio, exponentiated coefficients of Poisson regression
95% CI = 95 percent confidence interval

Figure S1: Spaghetti plot of weekly releases of all-cause mortality for four states in the same region: Connecticut reports deaths 4.8 weeks slower than average, and New York State (minus New York City), New Jersey, and Pennsylvania report deaths 2.5, 1.6, and 1.3 weeks faster than average.

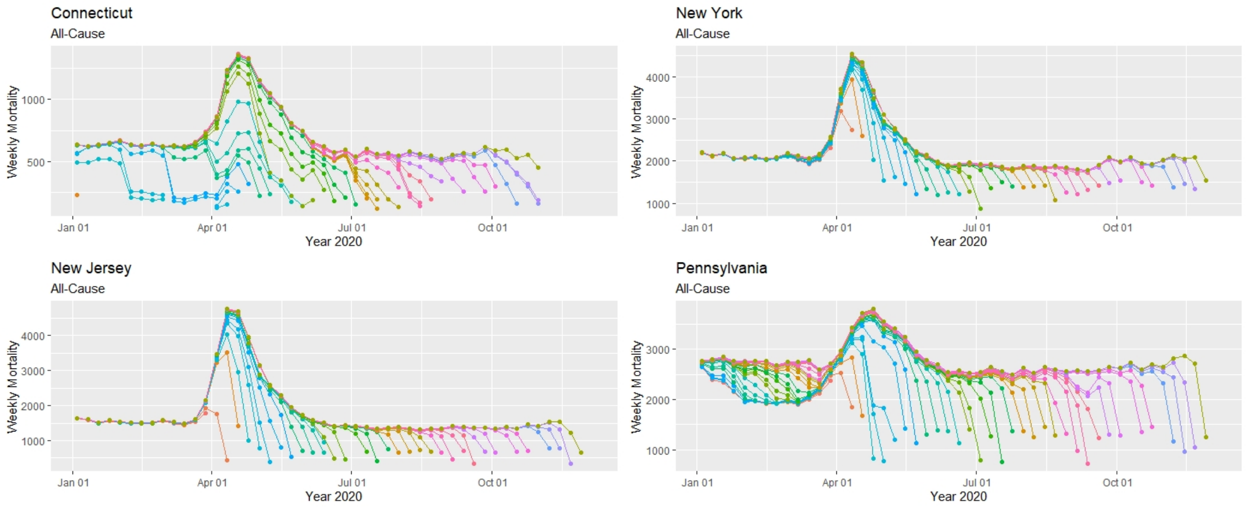


Each data point represents a provisional mortality count for that date. Each color line represents each of the 35 data releases between April 17, 2020 and December 4, 2020 in chromatic order; the chromatic order is used instead of a legend for each of the 35 colors.

Figure S2: Spaghetti plot of weekly releases of all-cause mortality for the three fastest states and three slowest states. Each data point represents a provisional mortality count for that date. Each color line represents each of the 35 data releases between April 17, 2020 and December 4, 2020 in chromatic order; the chromatic order is used instead of a legend for each of the 35 colors.

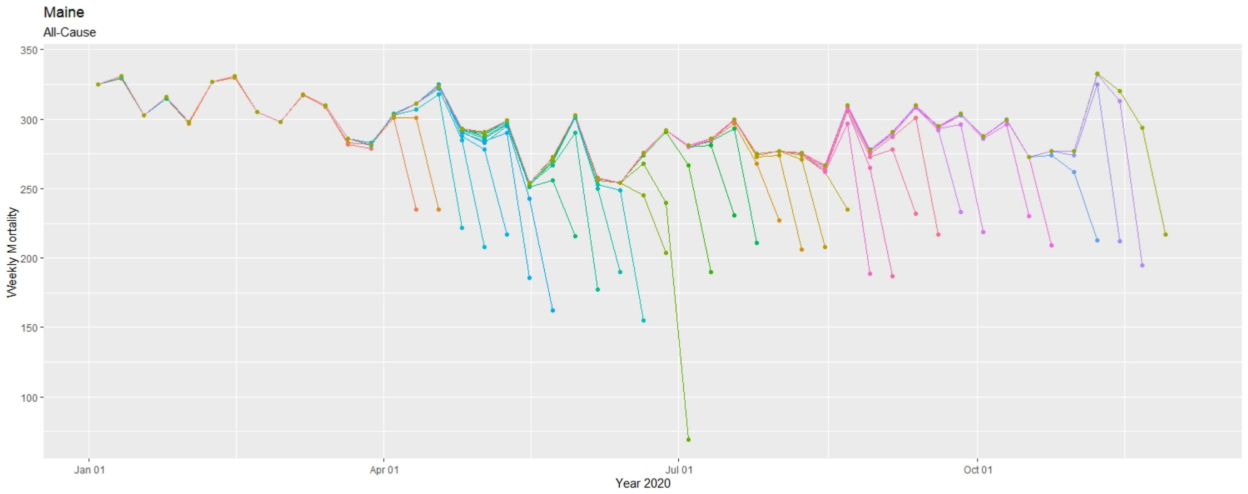
Maine
All-Cause
Weekly Mortality
350
300
250
200
150
100
Jan 01
Apr 01
Jul 01
Oct 01
Year 2020

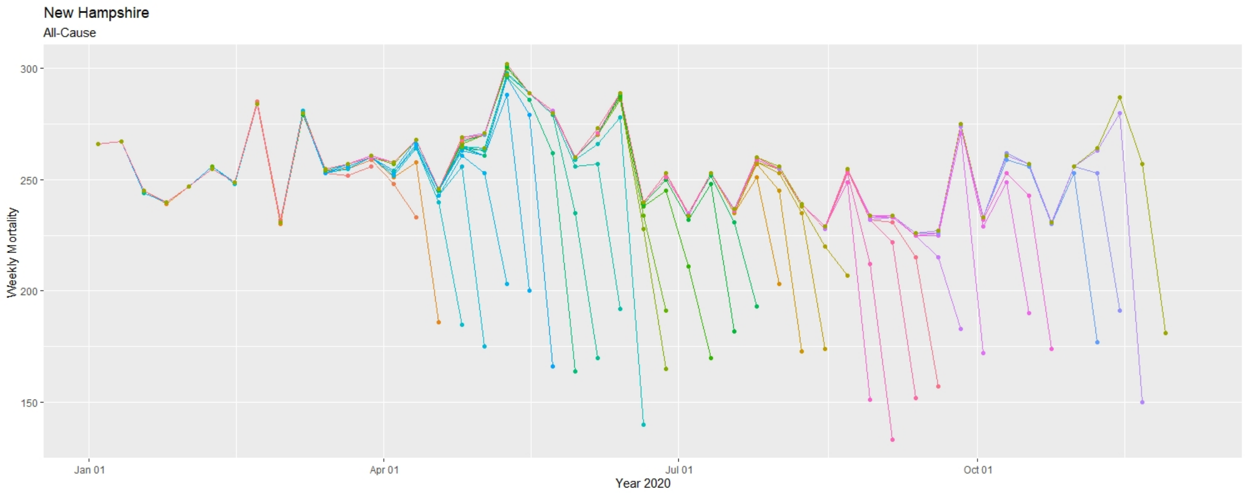
New Hampshire
All-Cause
Weekly Mortality
300
250
200
150
Jan 01
Apr 01
Jul 01
Oct 01
Year 2020

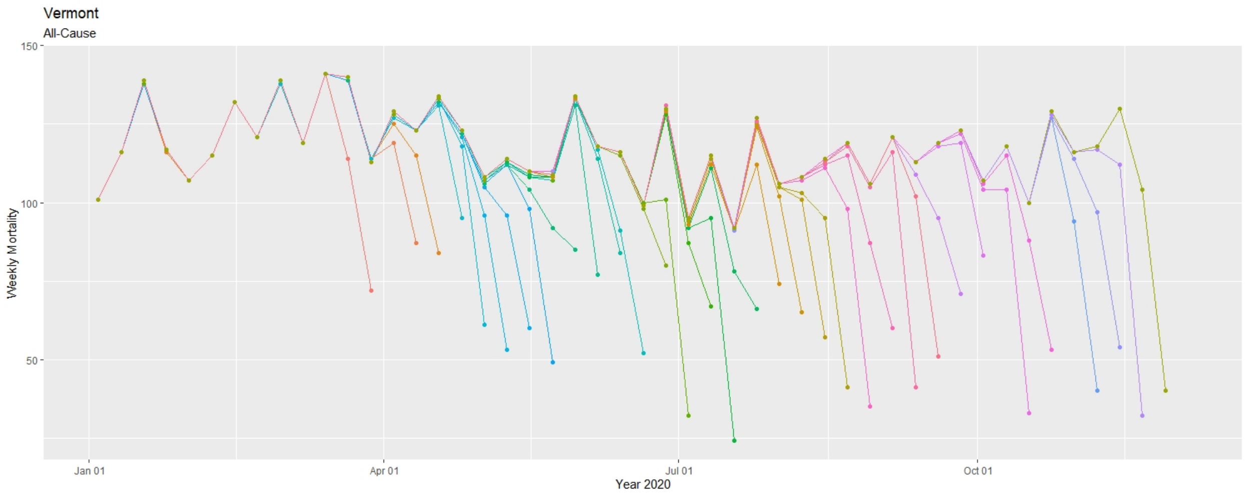
Vermont
All-Cause
Weekly Mortality
150
100
50
Jan 01
Apr 01
Jul 01
Oct 01
Year 2020

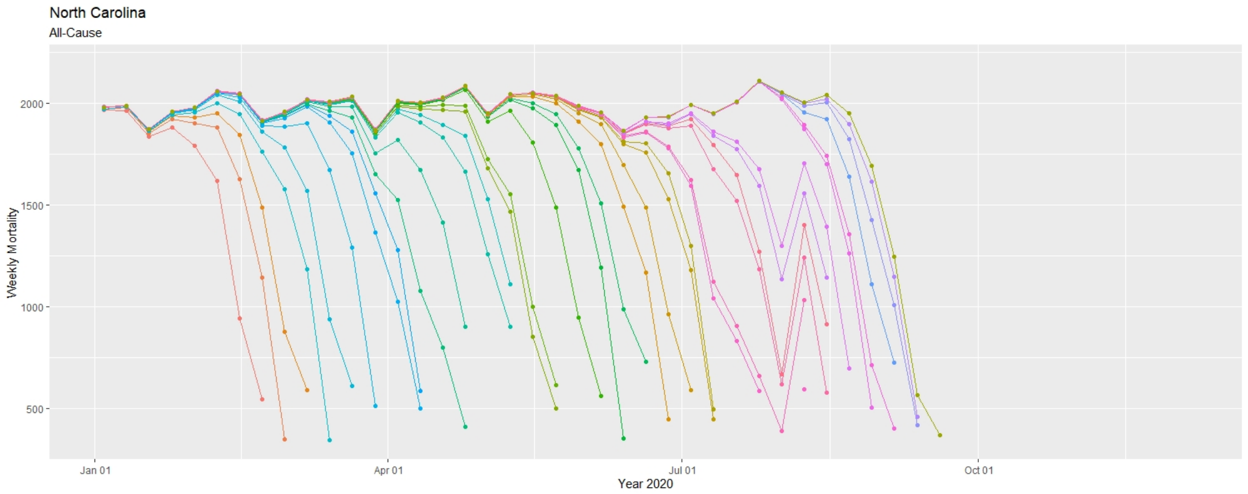
North Carolina
All-Cause
Weekly Mortality
2000
1500
1000
500
Jan 01
Apr 01
Jul 01
Oct 01
Year 2020

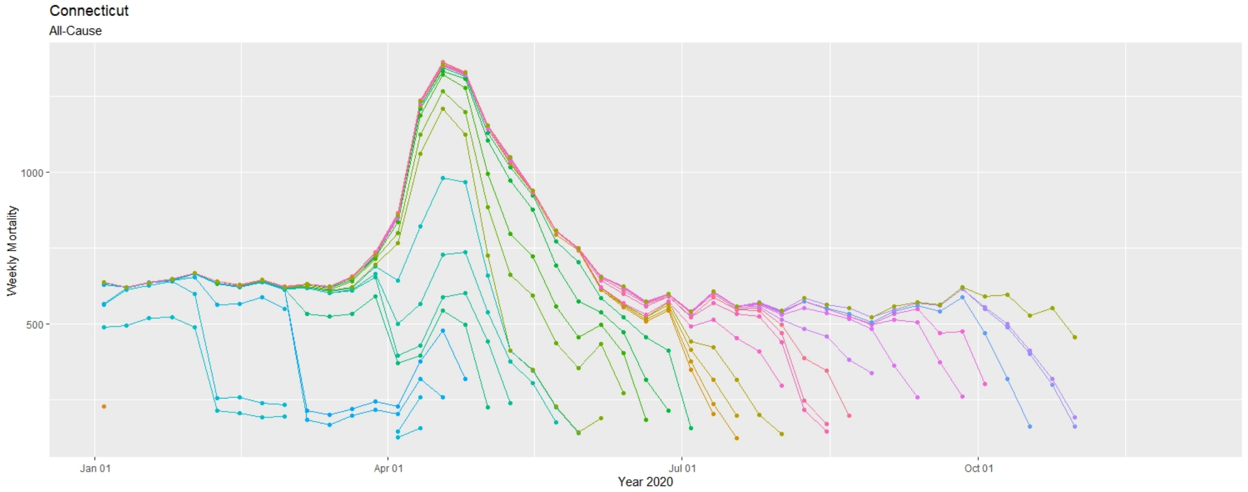
Connecticut
All-Cause
Weekly Mortality
1000
500
Jan 01
Apr 01
Jul 01
Oct 01
Year 2020

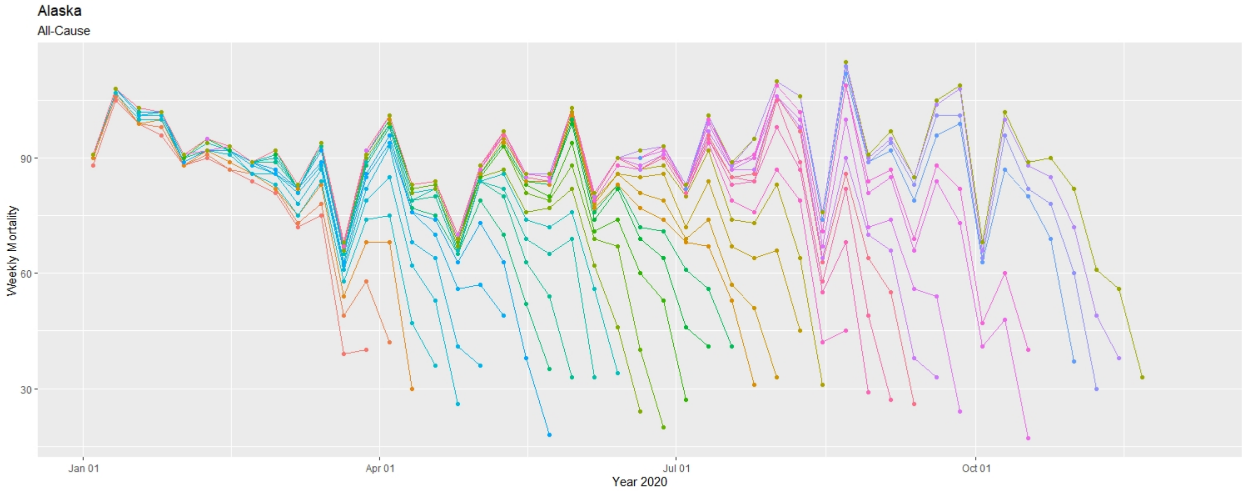
Alaska
All-Cause
Weekly Mortality
90
60
30
Jan 01
Apr 01
Jul 01
Oct 01
Year 2020

Figure S3: Weeks until all-cause mortality counts are complete for April 3-September 5, 2020, adjusted for deaths per population. The red line shows the mean delay.

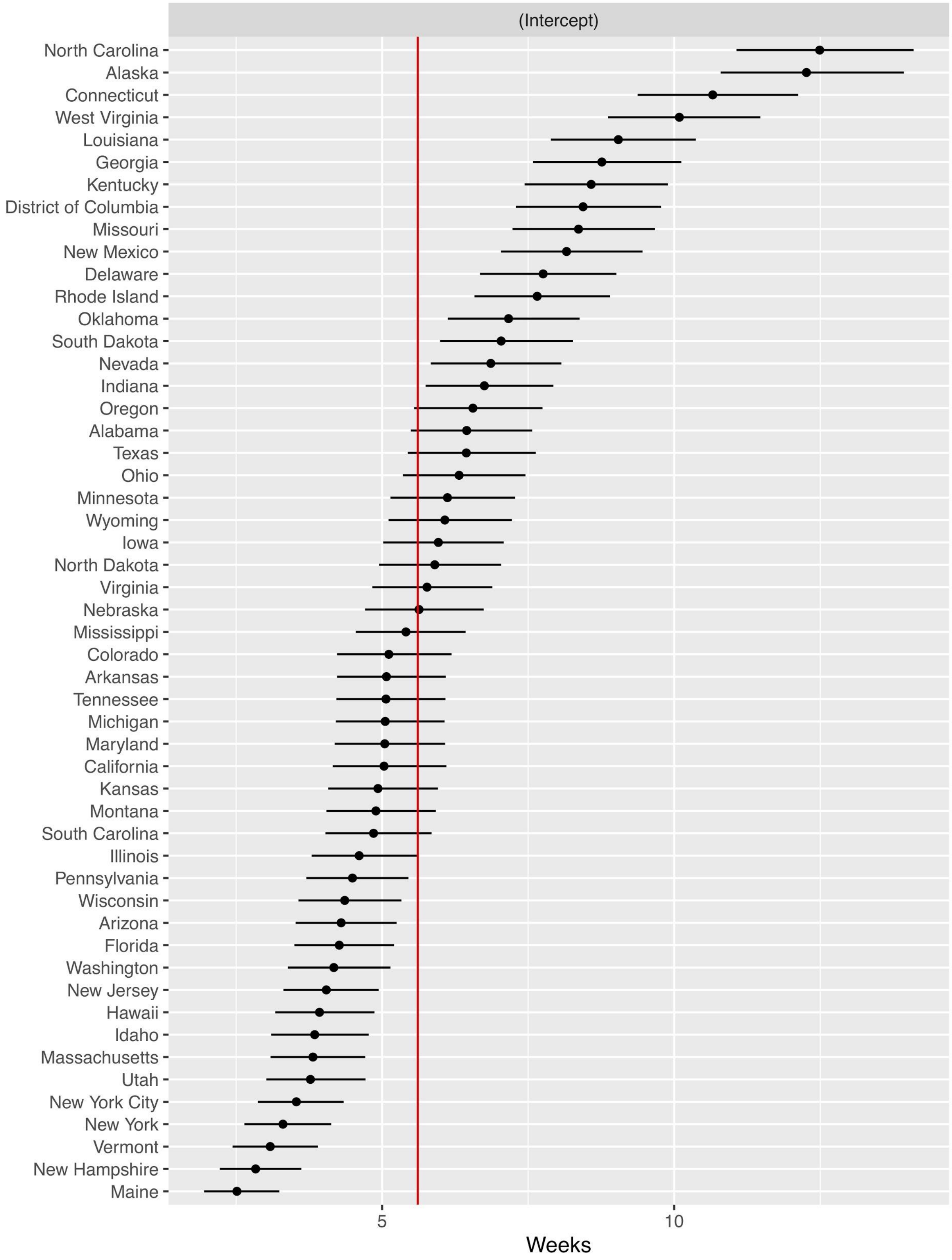

Figure S4: Spaghetti plot of weekly releases of all-cause mortality for all 50 states. Each data point represents a provisional mortality count for that date. Each color line represents each of the 35 data releases between April 17, 2020 and December 4, 2020 in chromatic order; the chromatic order is used instead of a legend for each of the 35 colors.

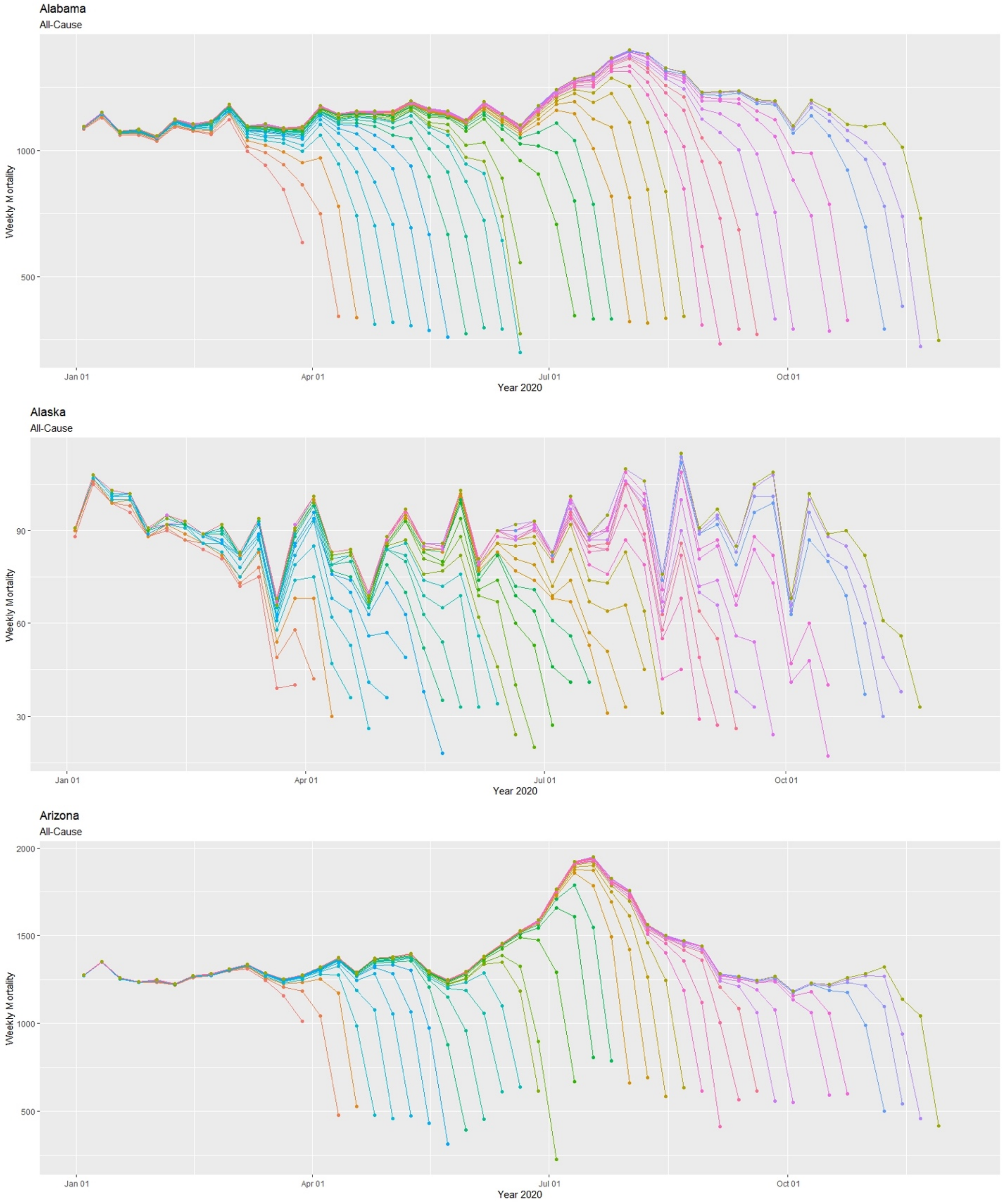

Arkansas
All-Cause

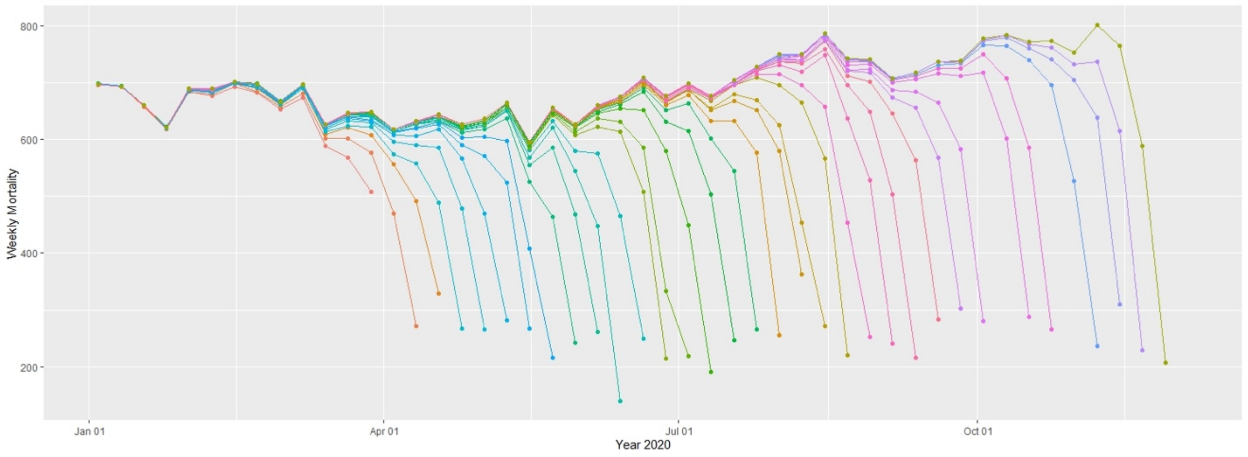


California
All-Cause

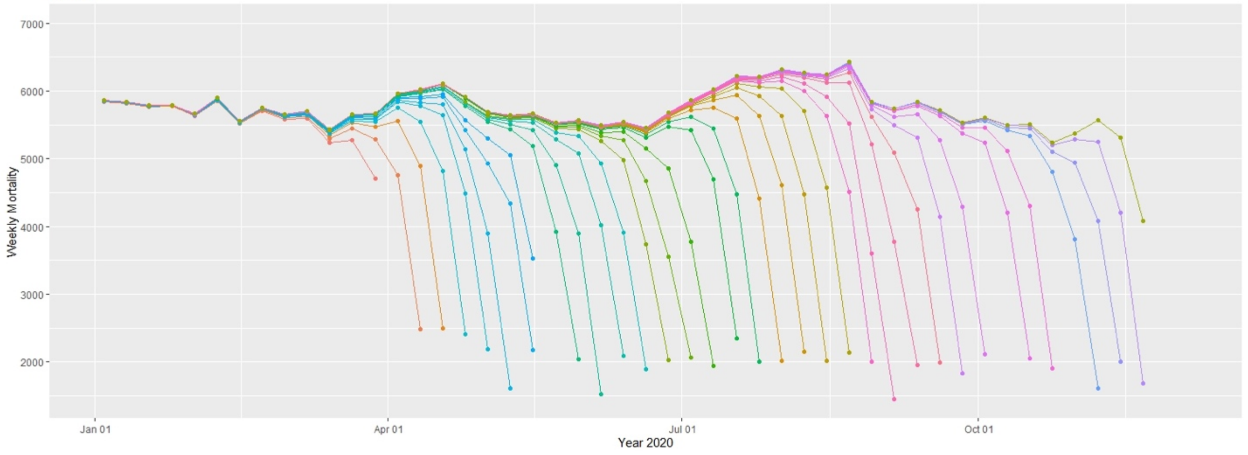


Colorado
All-Cause

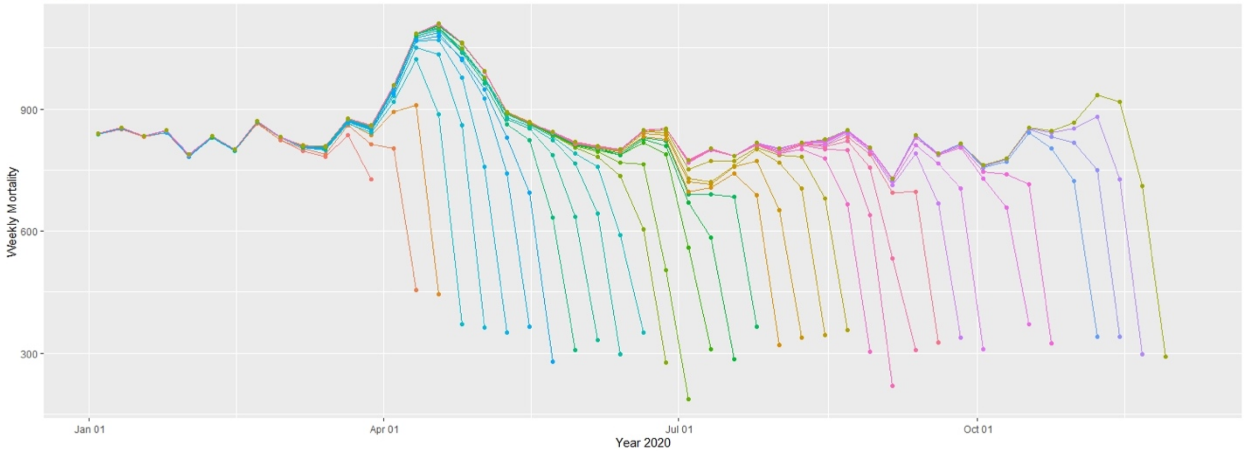

Connecticut

All-Cause

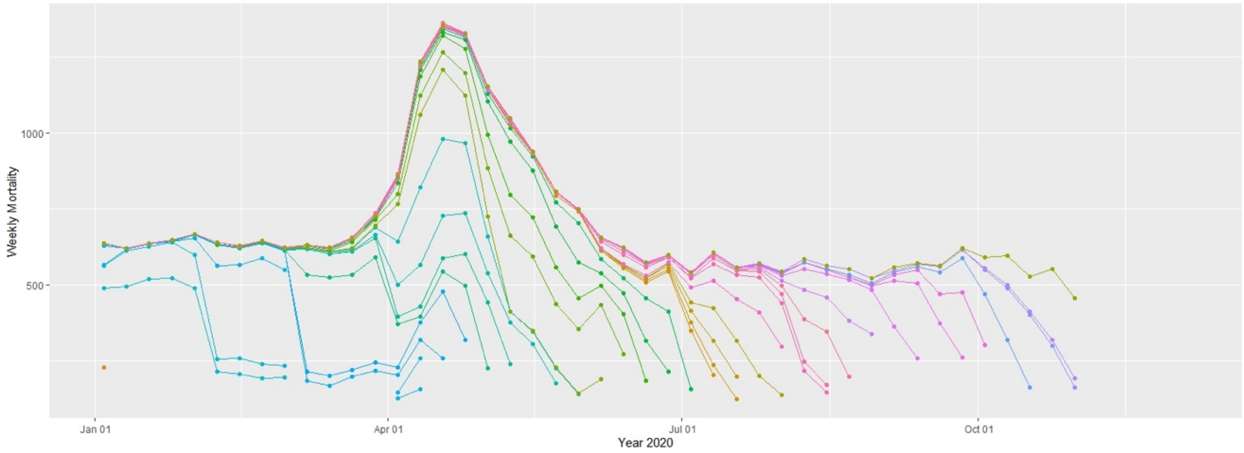


Delaware

All-Cause

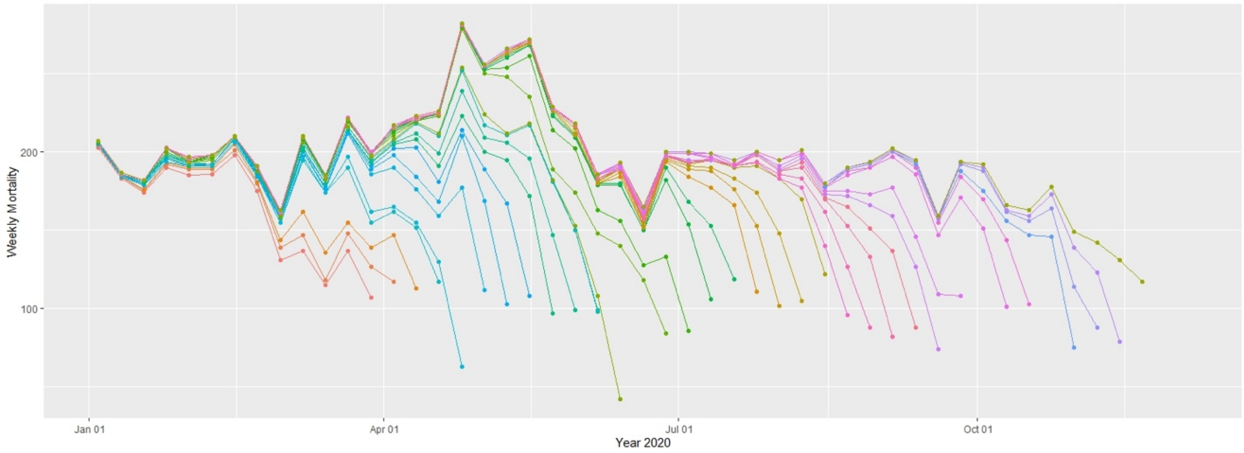


District of Columbia

All-Cause

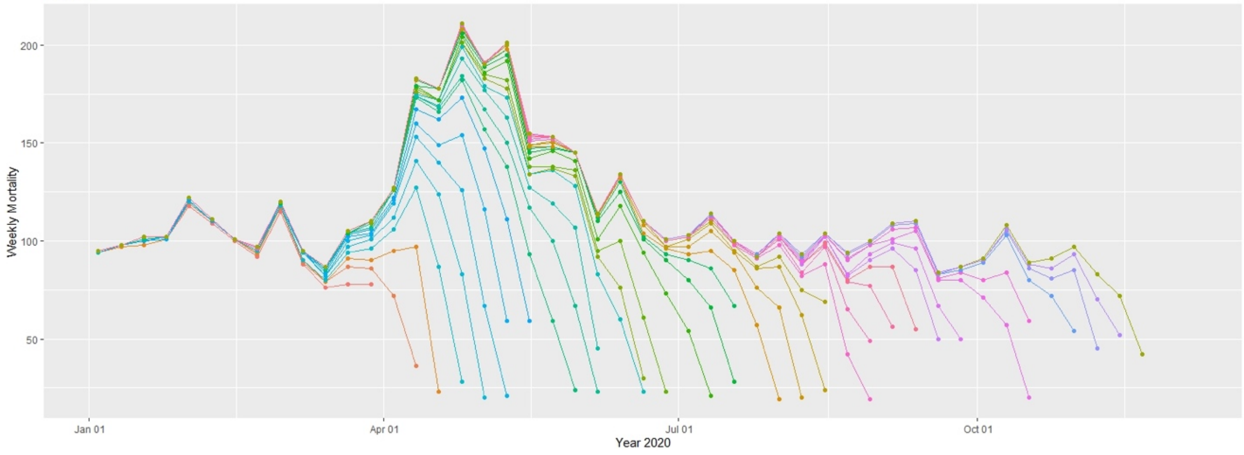

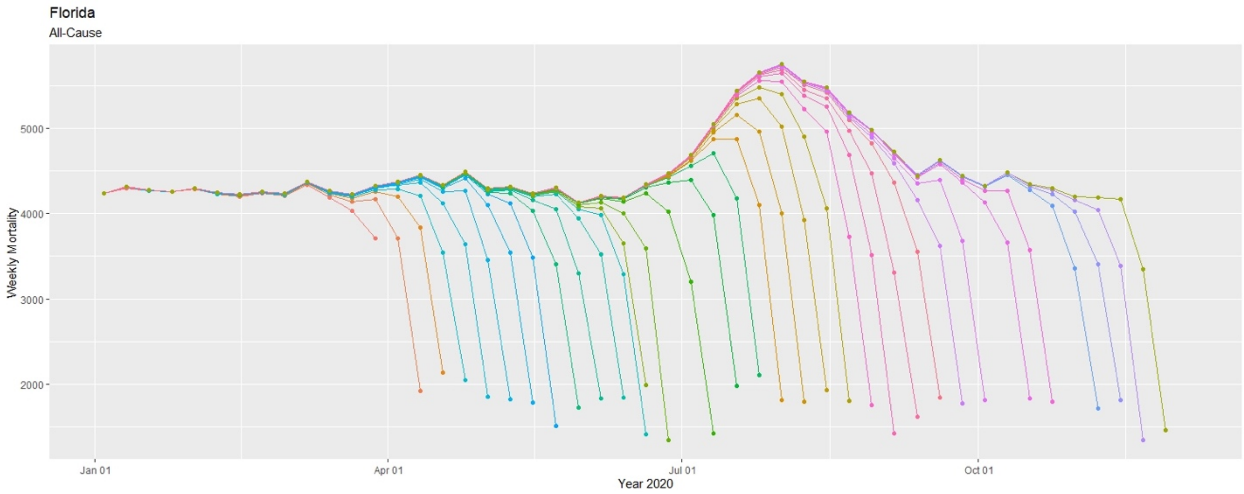
Florida
All-Cause
Weekly Mortality
5000
4000
3000
2000
Jan 01
Apr 01
Jul 01
Oct 01
Year 2020

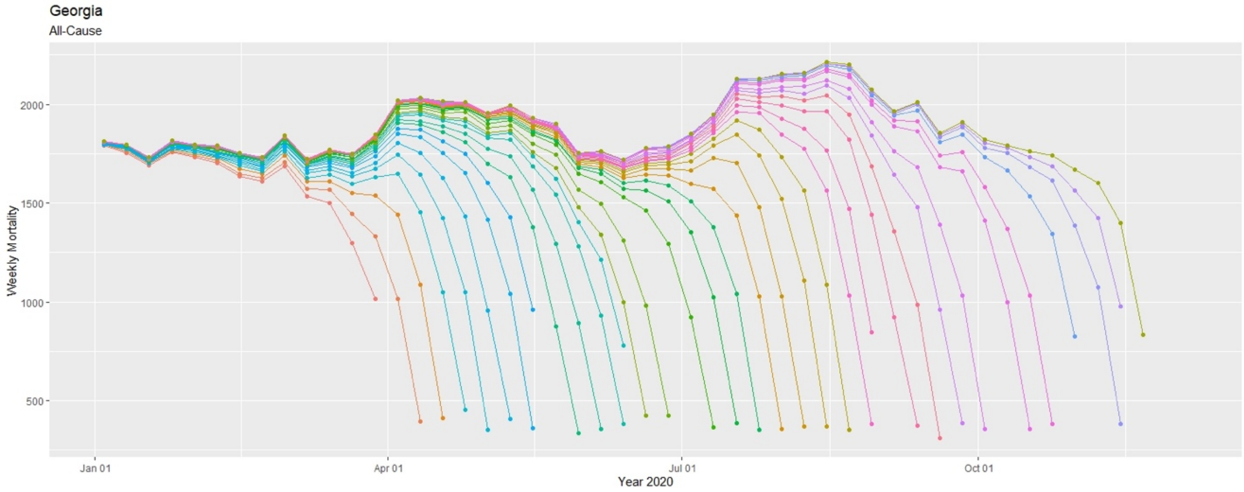
Georgia
All-Cause
Weekly Mortality
2000
1500
1000
500
Jan 01
Apr 01
Jul 01
Oct 01
Year 2020

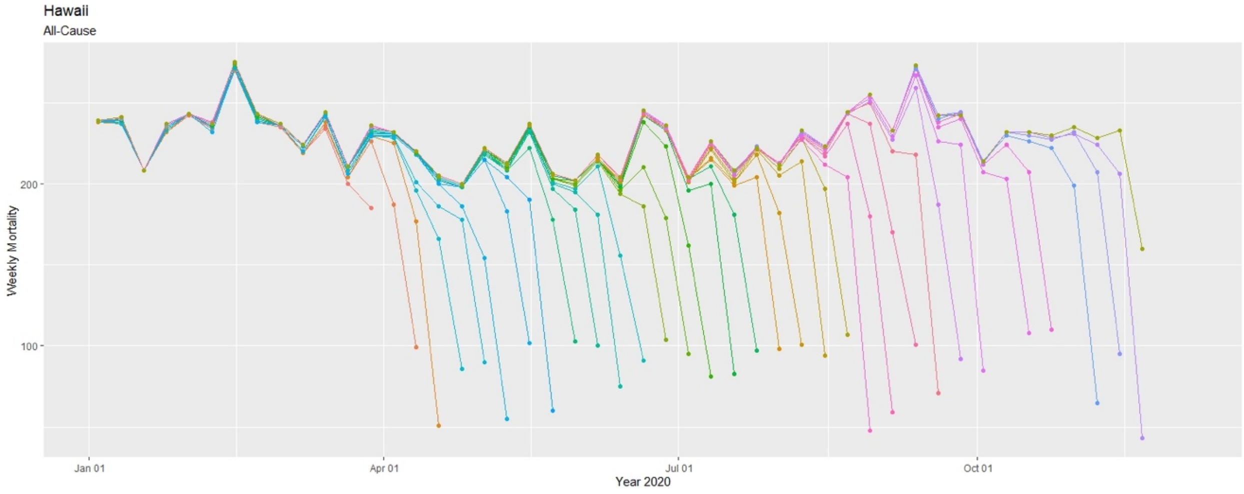
Hawaii
All-Cause
Weekly Mortality
200
100
Jan 01
Apr 01
Jul 01
Oct 01
Year 2020

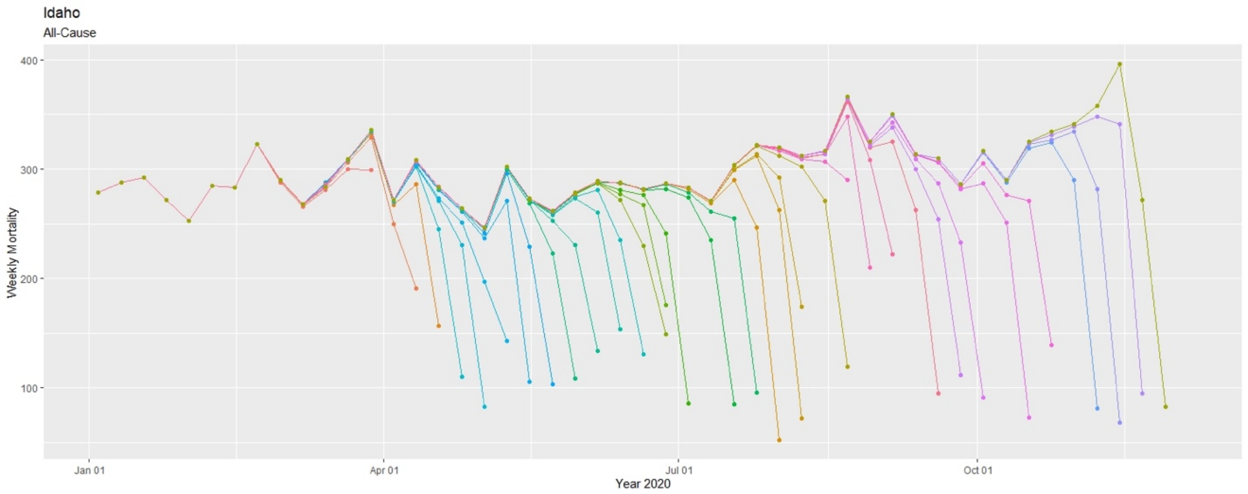

Idaho
All-Cause
Weekly Mortality
400
300
200
100
Jan 01
Apr 01
Jul 01
Oct 01
Year 2020


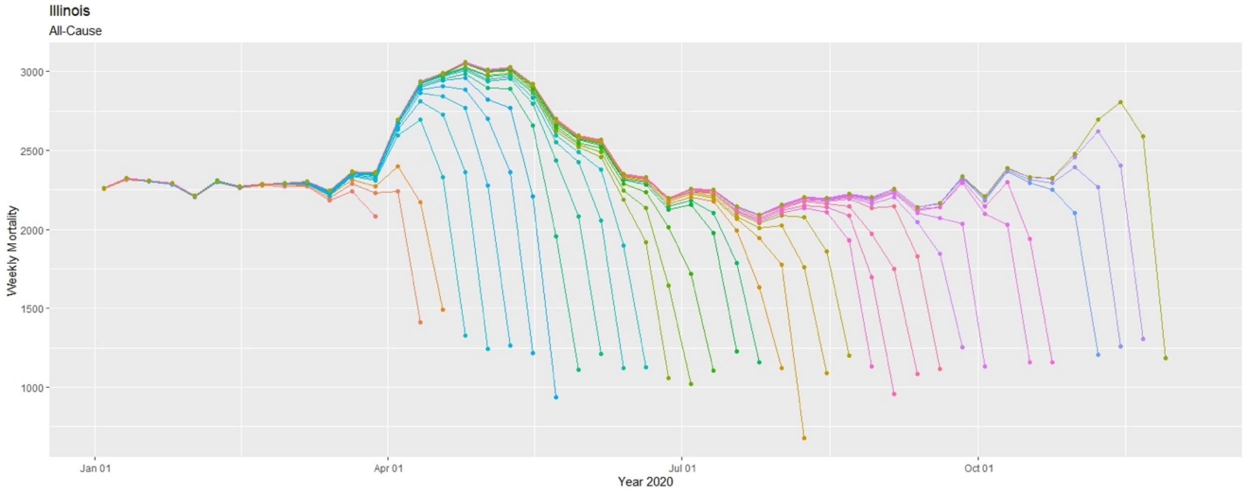

Illinois
All-Cause
Weekly Mortality
3000
2500
2000
1500
1000
Jan 01
Apr 01
Jul 01
Oct 01
Year 2020


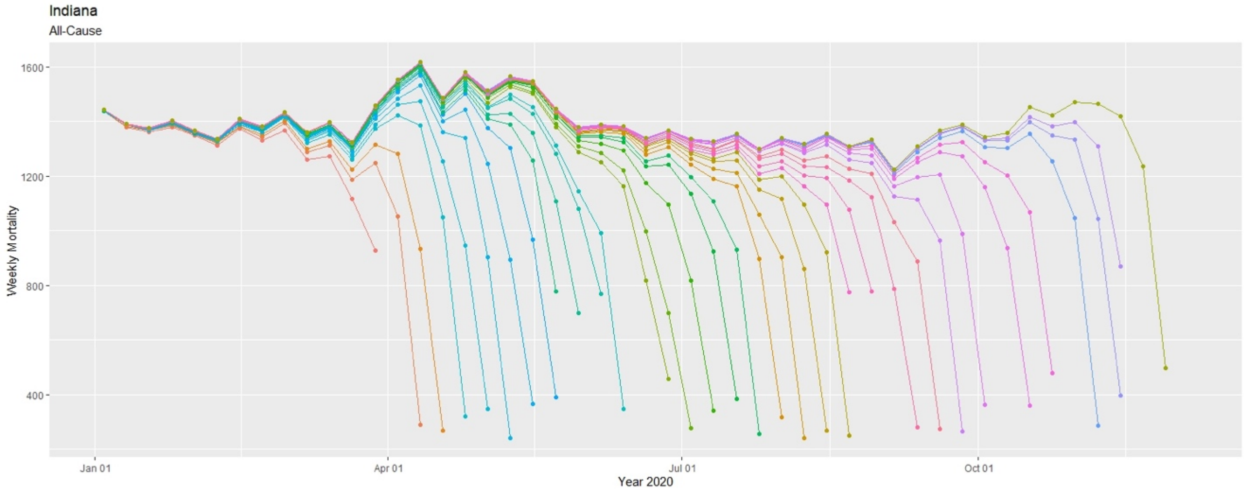

Indiana
All-Cause
Weekly Mortality
1600
1200
800
400
Jan 01
Apr 01
Jul 01
Oct 01
Year 2020

Iowa
All-Cause

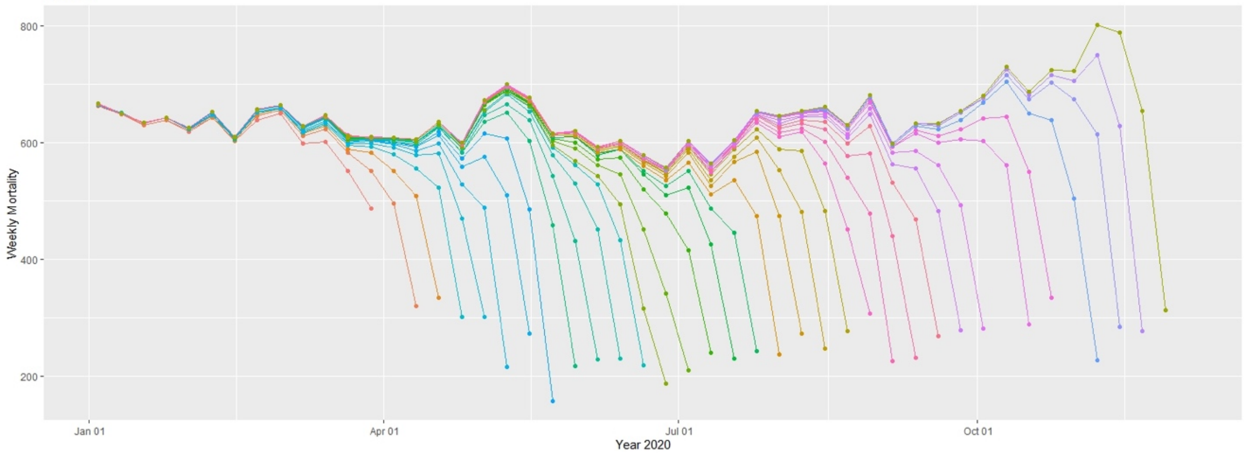


Kansas
All-Cause

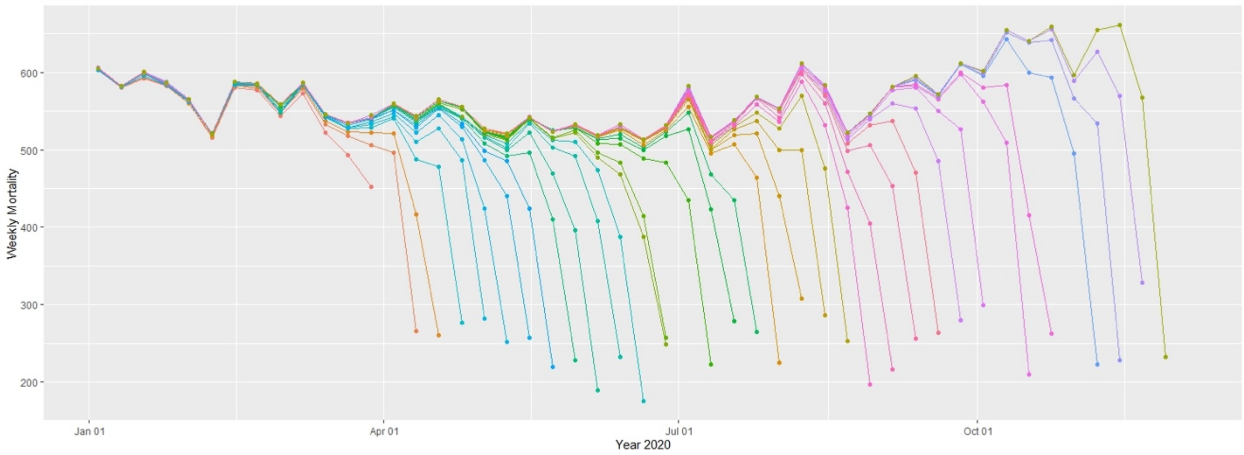


Kentucky
All-Cause

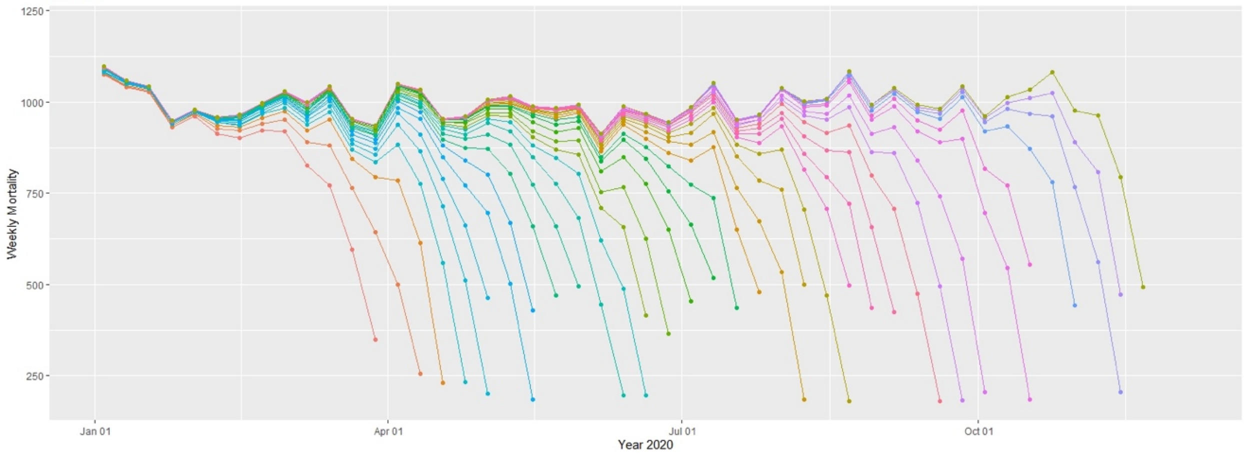

Louisiana

All-Cause

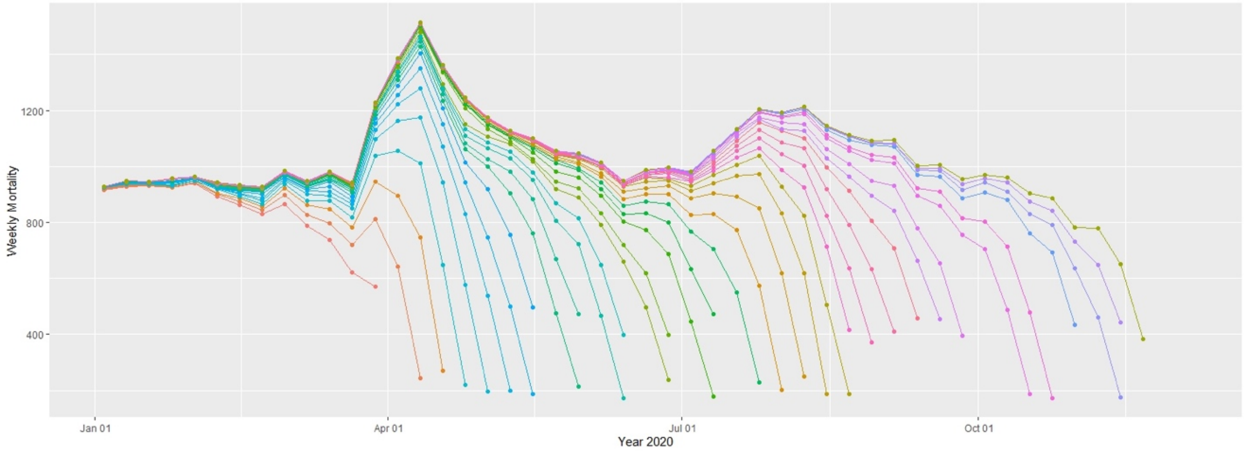


Maine

All-Cause

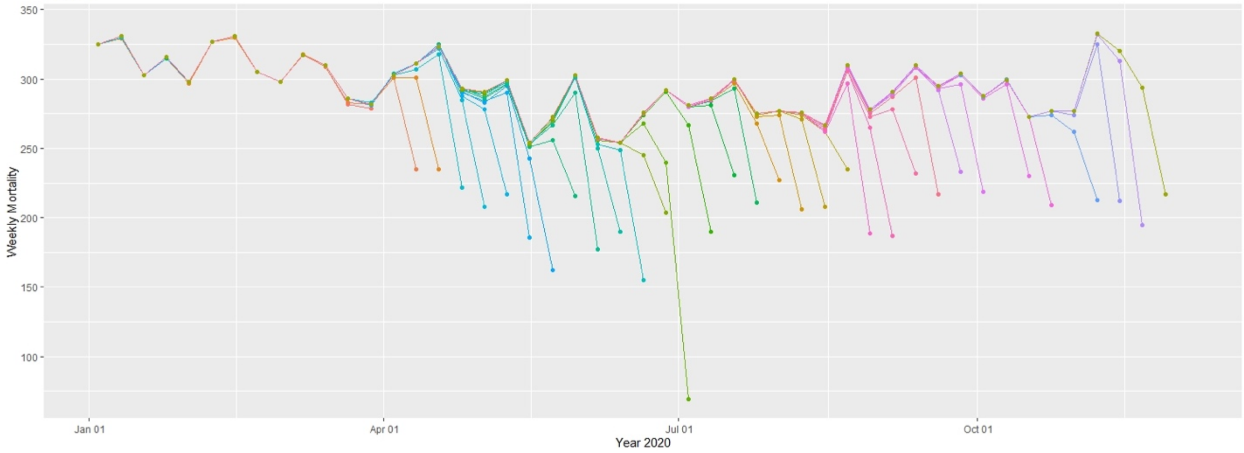


Maryland

All-Cause

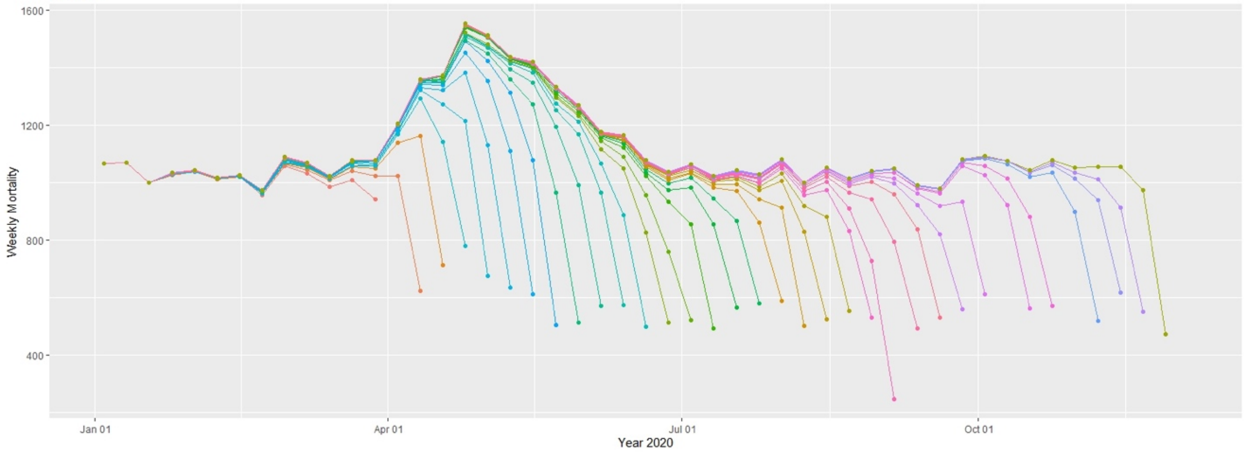

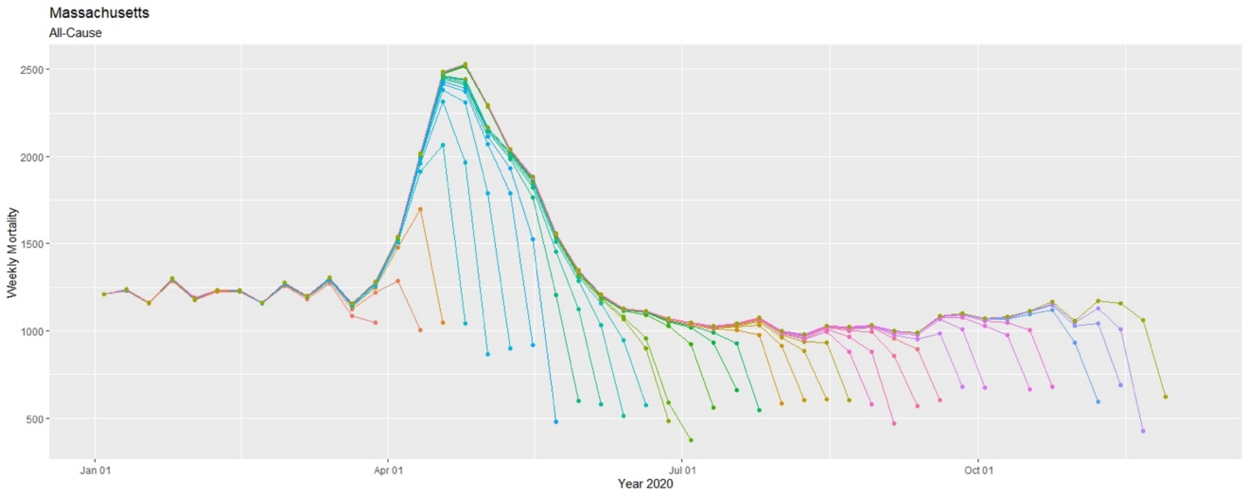
Massachusetts
All-Cause
Weekly Mortality
2500
2000
1500
1000
500
Jan 01
Apr 01
Jul 01
Oct 01
Year 2020

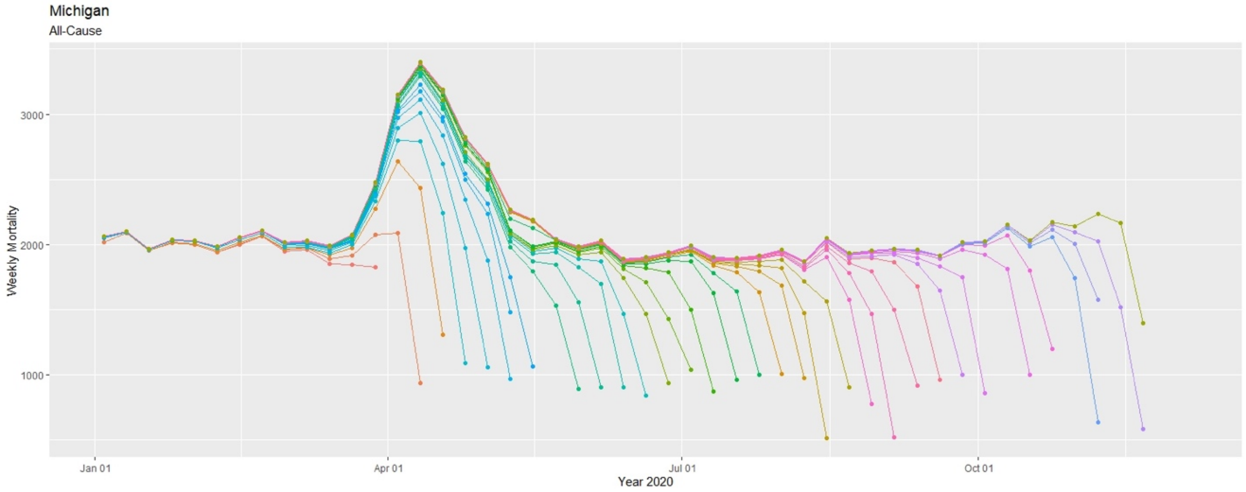
Michigan
All-Cause
Weekly Mortality
3000
2000
1000
Jan 01
Apr 01
Jul 01
Oct 01
Year 2020

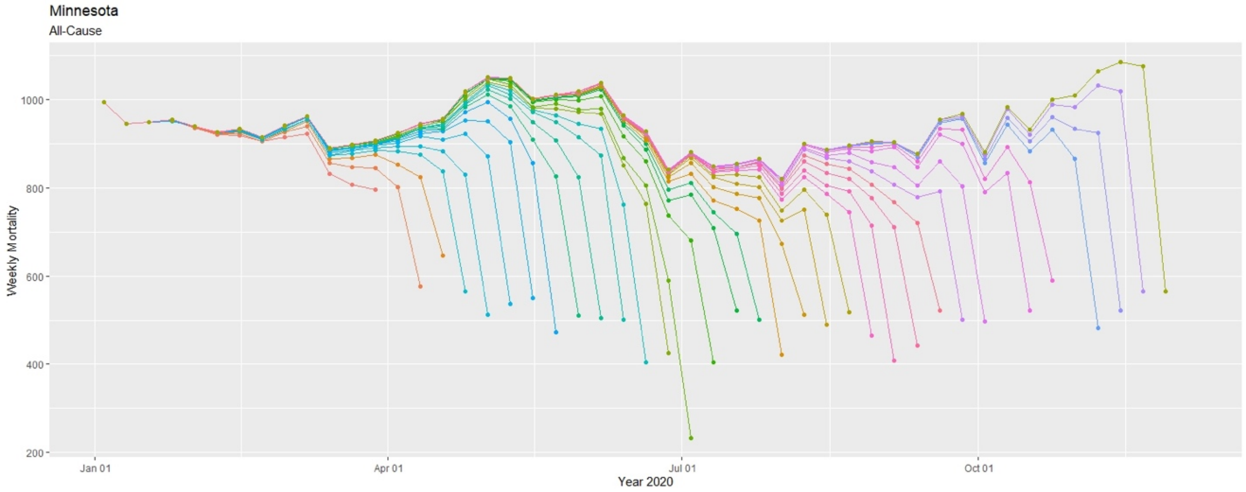
Minnesota
All-Cause
Weekly Mortality
1000
800
600
400
200
Jan 01
Apr 01
Jul 01
Oct 01
Year 2020

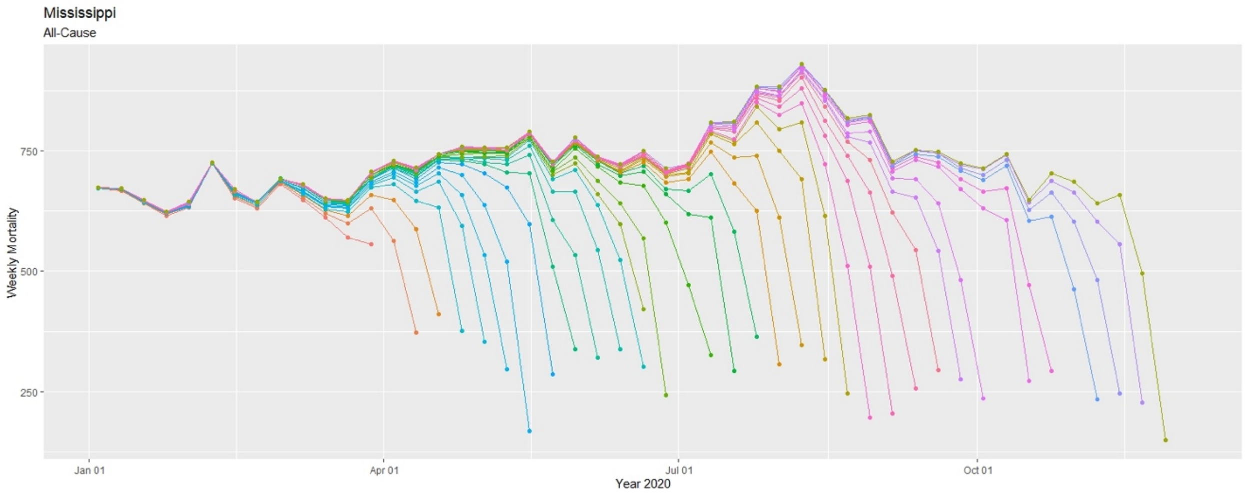
Mississippi
All-Cause
Weekly Mortality
750
500
250
Jan 01
Apr 01
Jul 01
Oct 01
Year 2020

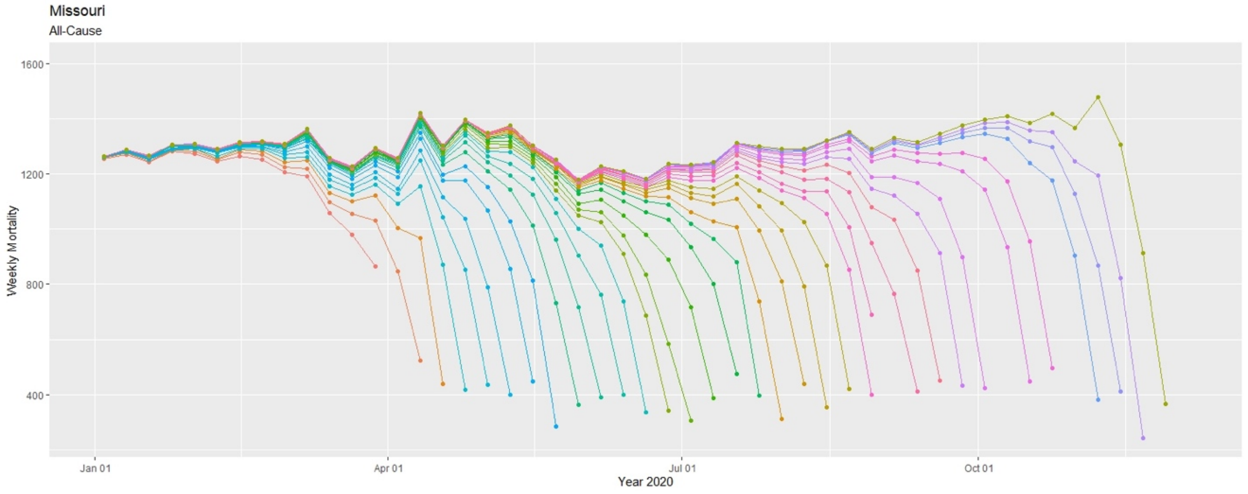
Missouri
All-Cause
Weekly Mortality
1600
1200
800
400
Jan 01
Apr 01
Jul 01
Oct 01
Year 2020

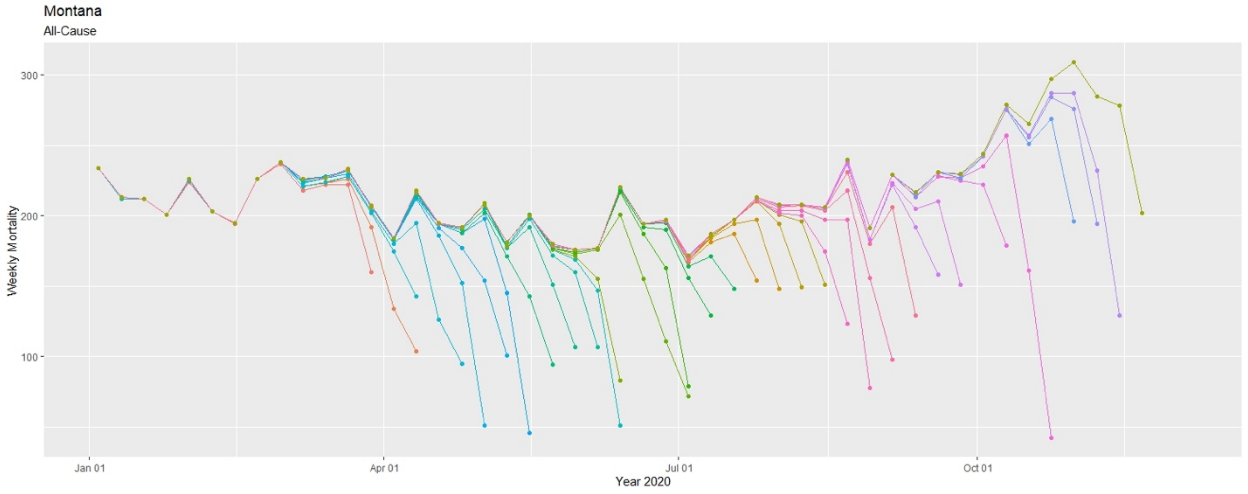
Montana
All-Cause
Weekly Mortality
300
200
100
Jan 01
Apr 01
Jul 01
Oct 01
Year 2020

Nebraska
All-Cause

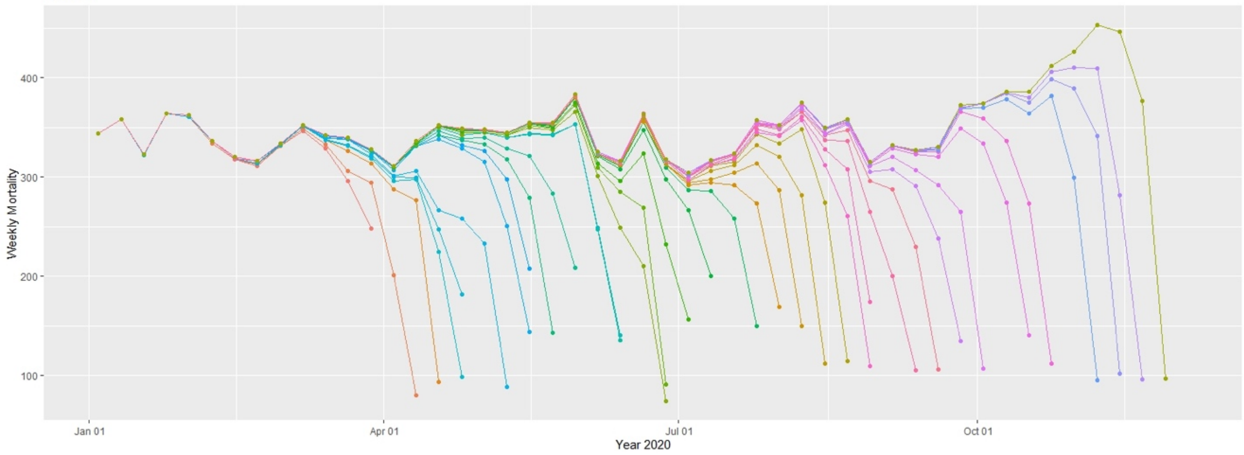


Nevada
All-Cause

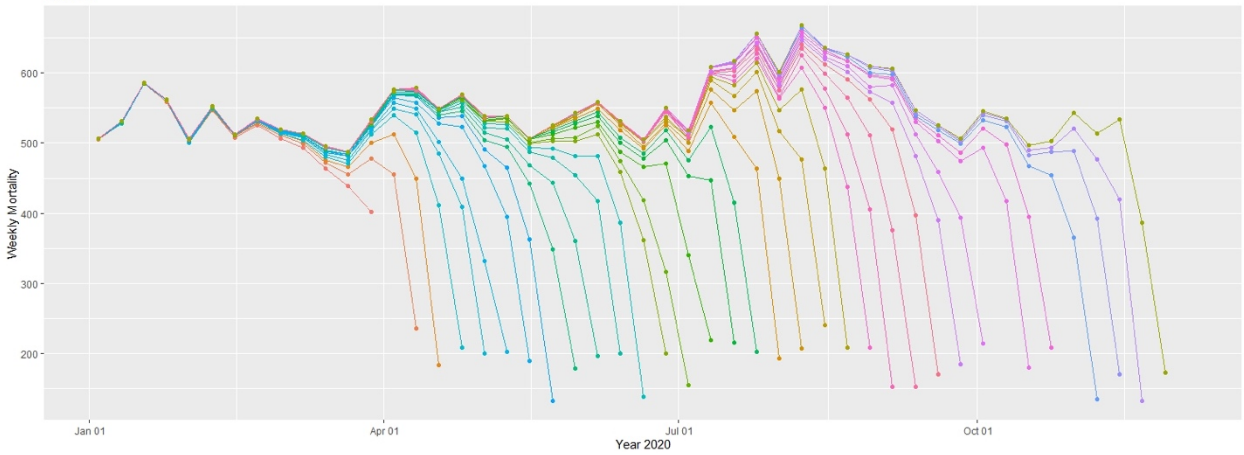


New Hampshire
All-Cause

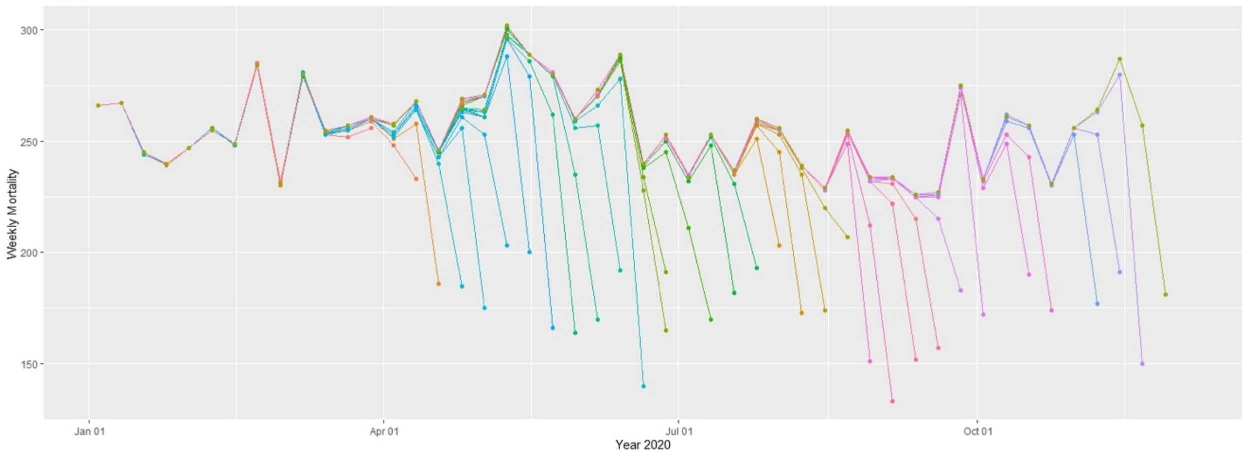

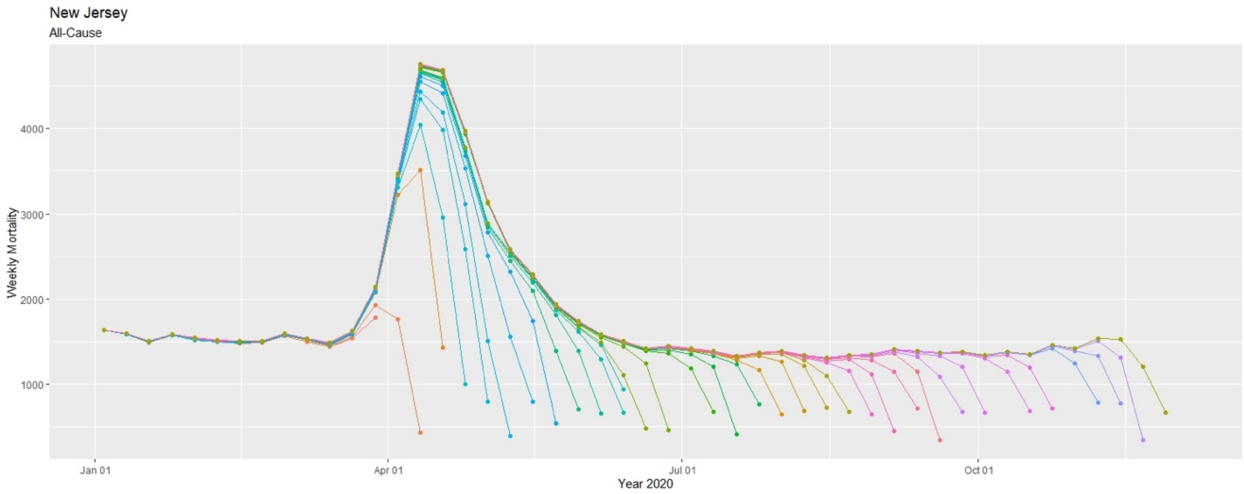
New Jersey
All-Cause
Weekly Mortality
4000
3000
2000
1000
Jan 01
Apr 01
Jul 01
Oct 01
Year 2020

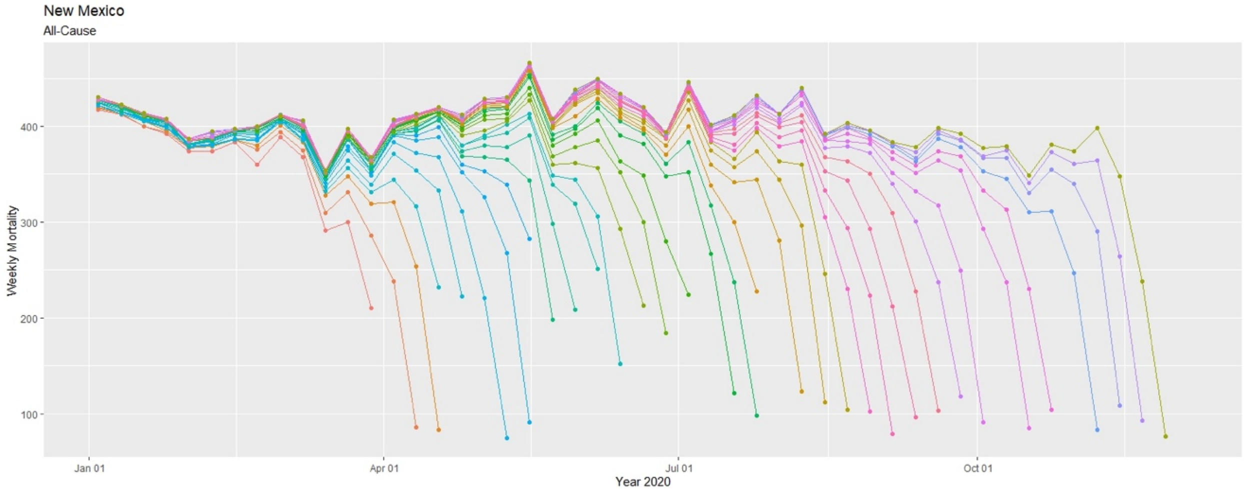
New Mexico
All-Cause
Weekly Mortality
400
300
200
100
Jan 01
Apr 01
Jul 01
Oct 01
Year 2020

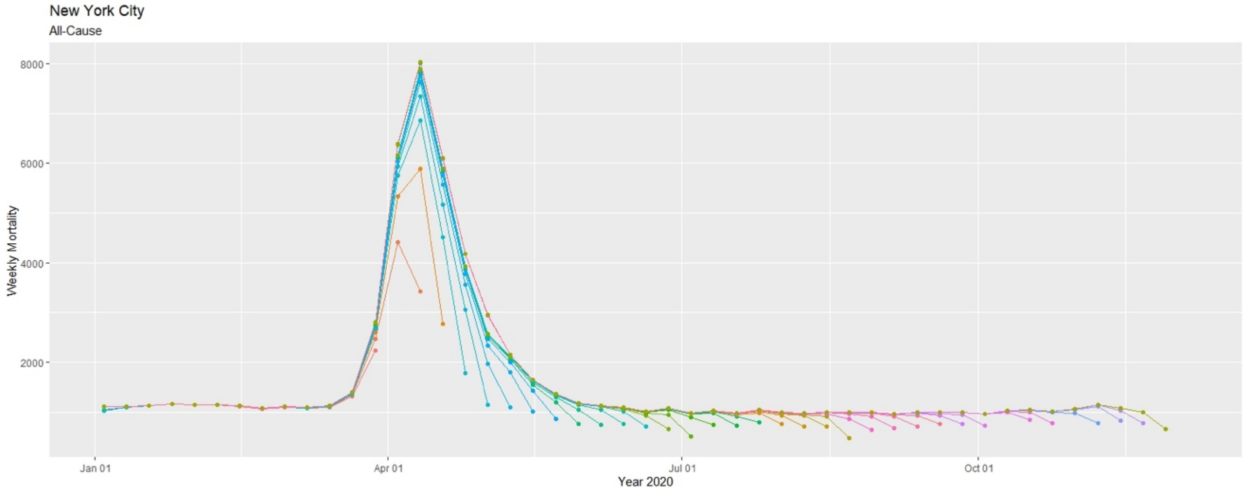
New York City
All-Cause
Weekly Mortality
8000
6000
4000
2000
Jan 01
Apr 01
Jul 01
Oct 01
Year 2020

New York
All-Cause

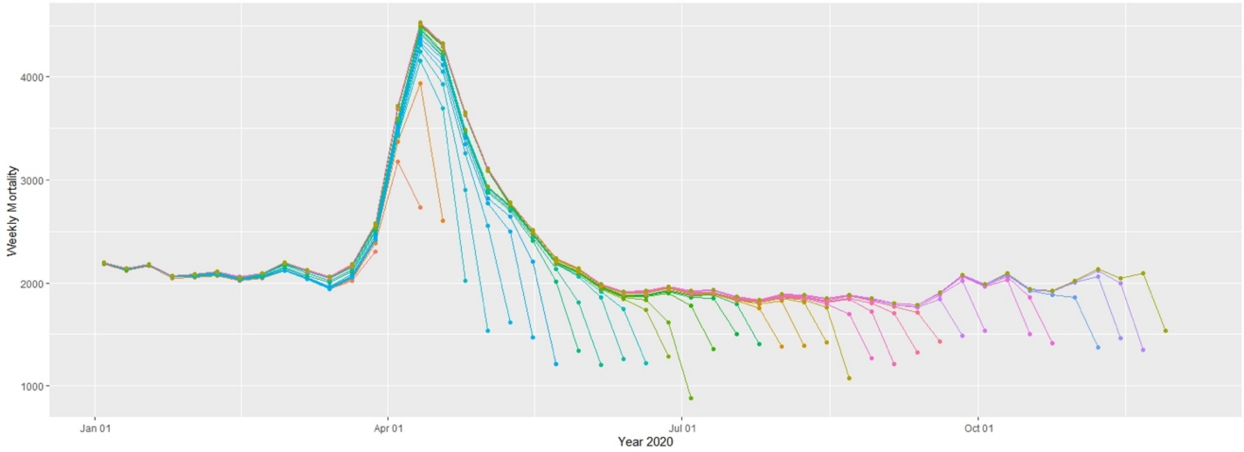


North Carolina
All-Cause

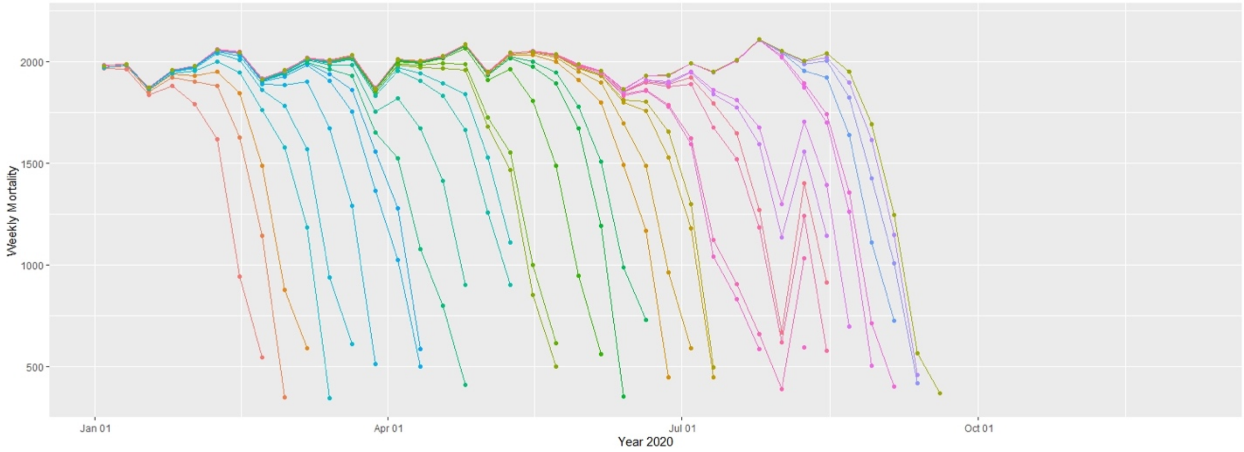


North Dakota
All-Cause

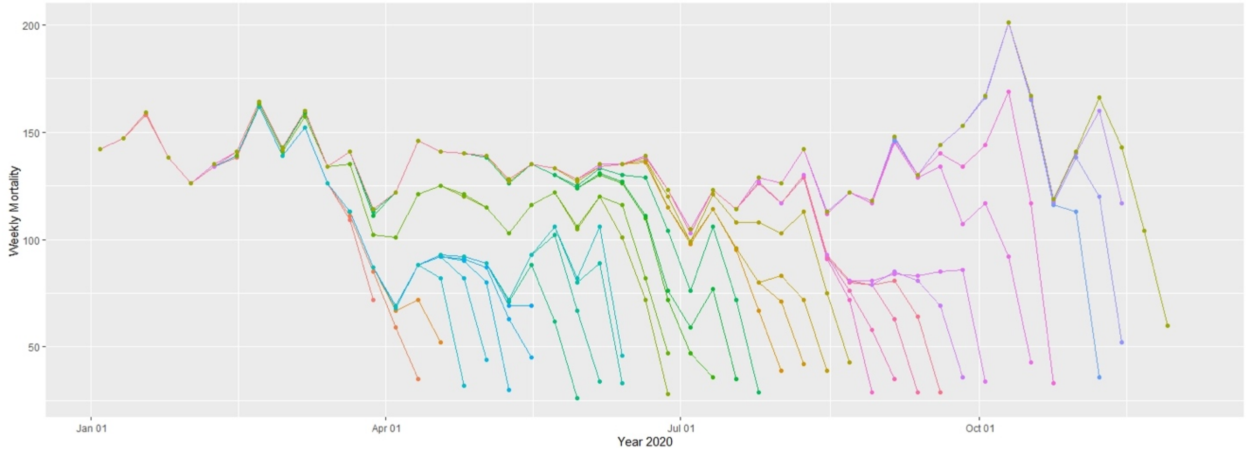

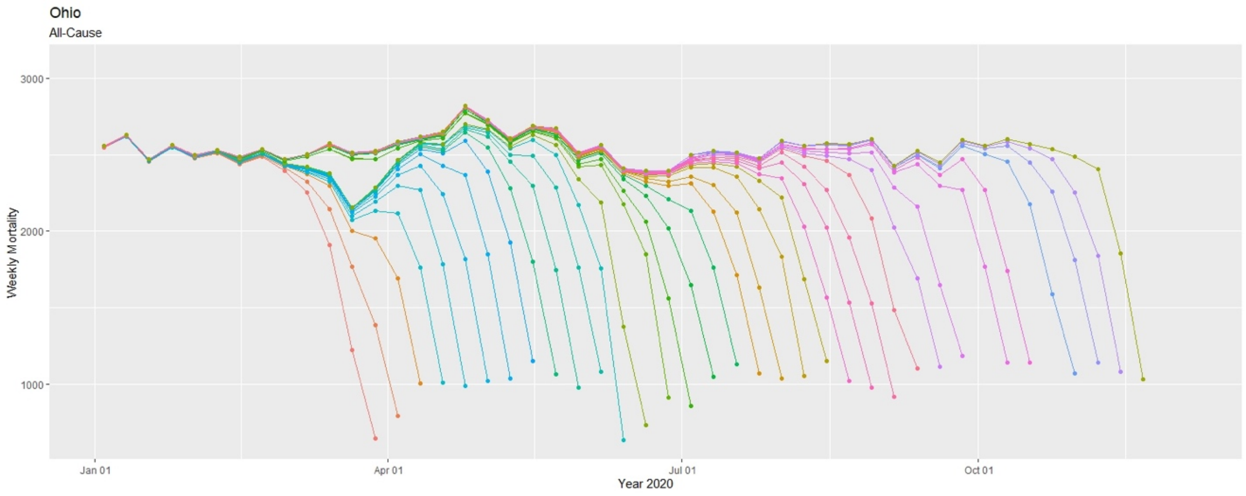
Ohio
All-Cause
Weekly Mortality
3000
2000
1000
Jan 01
Apr 01
Jul 01
Oct 01
Year 2020

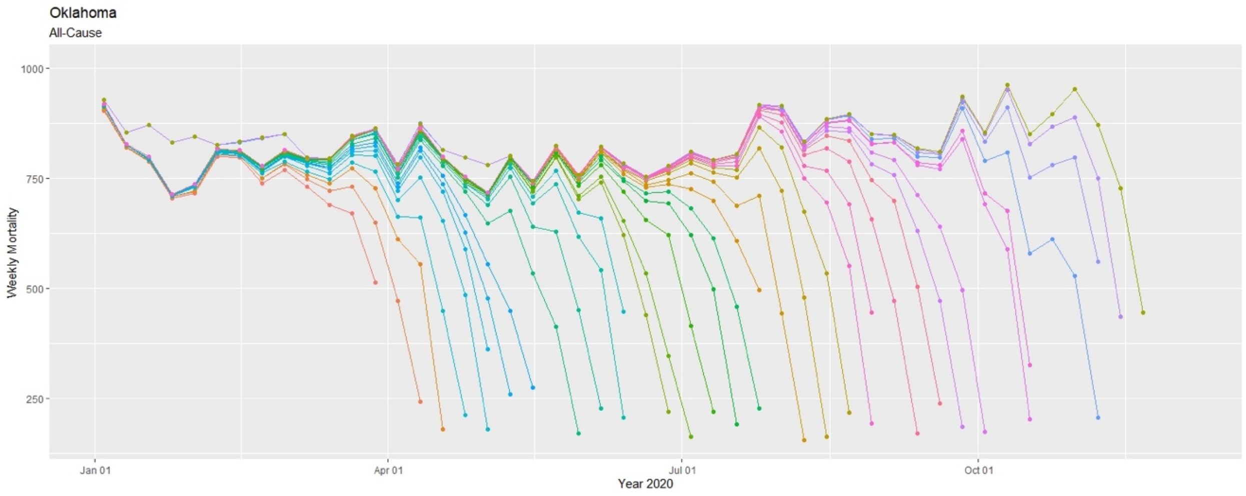
Oklahoma
All-Cause
Weekly Mortality
1000
750
500
250
Jan 01
Apr 01
Jul 01
Oct 01
Year 2020

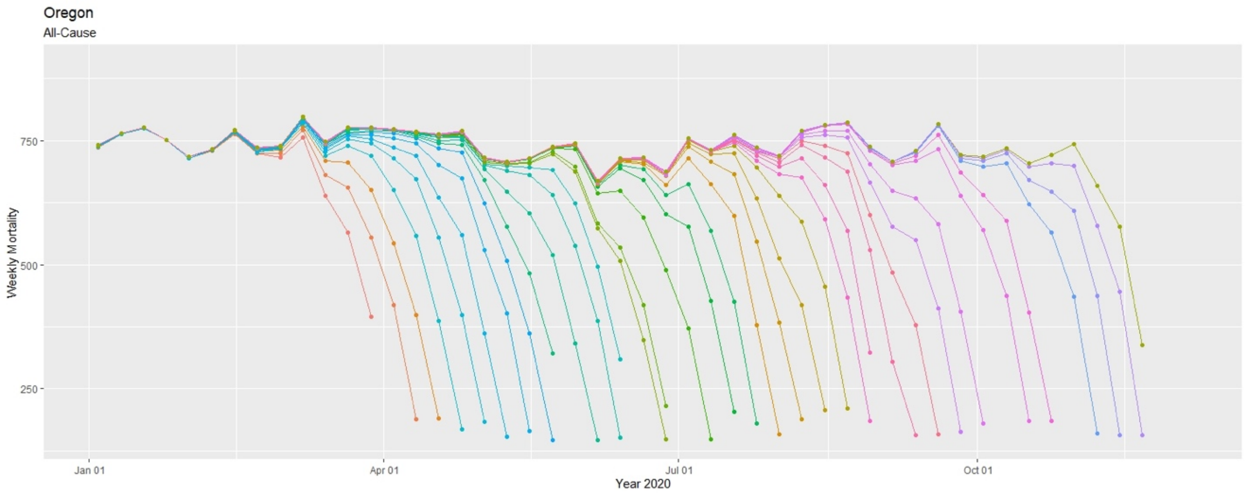
Oregon
All-Cause
Weekly Mortality
750
500
250
Jan 01
Apr 01
Jul 01
Oct 01
Year 2020

Pennsylvania
All-Cause

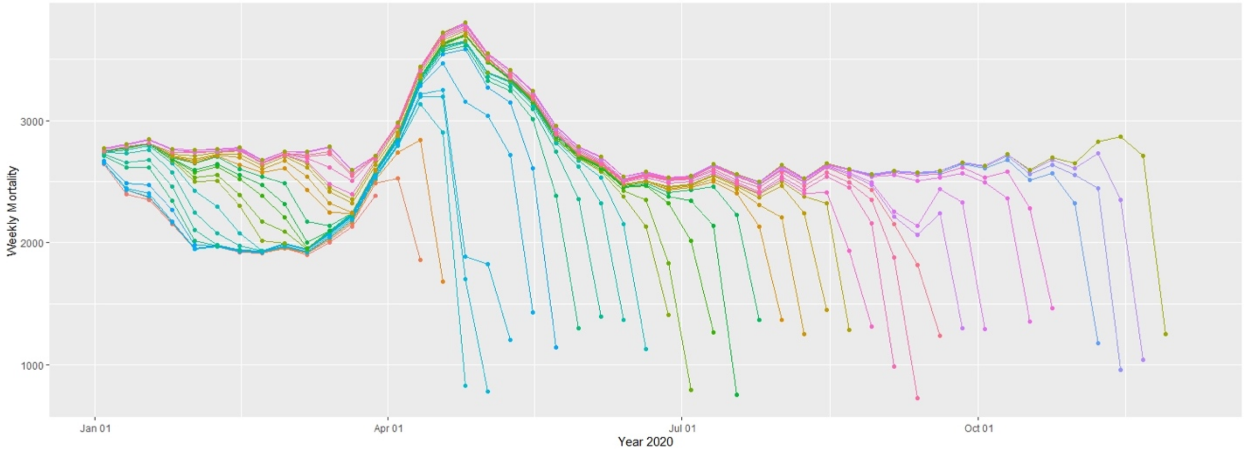


Rhode Island
All-Cause

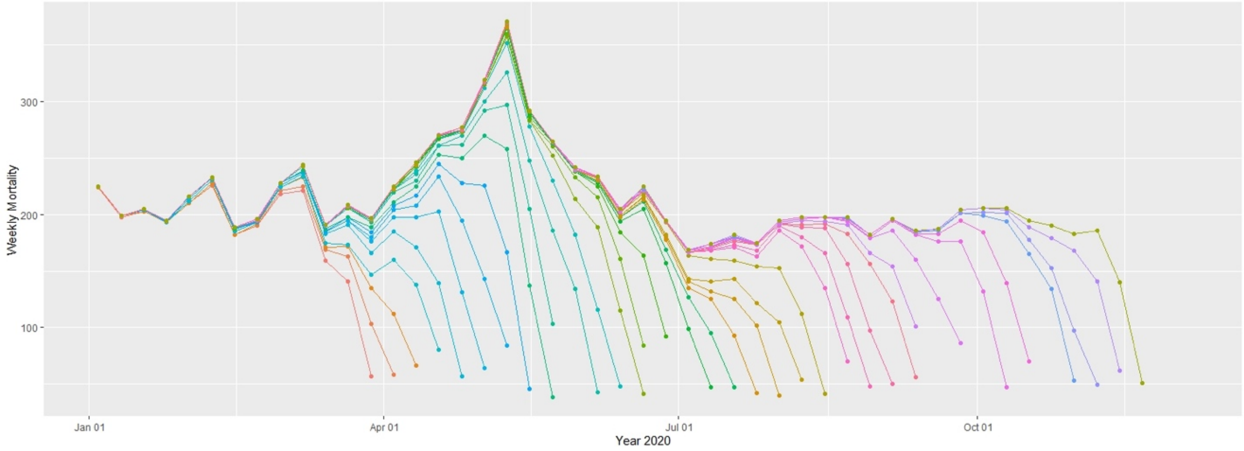


South Carolina
All-Cause

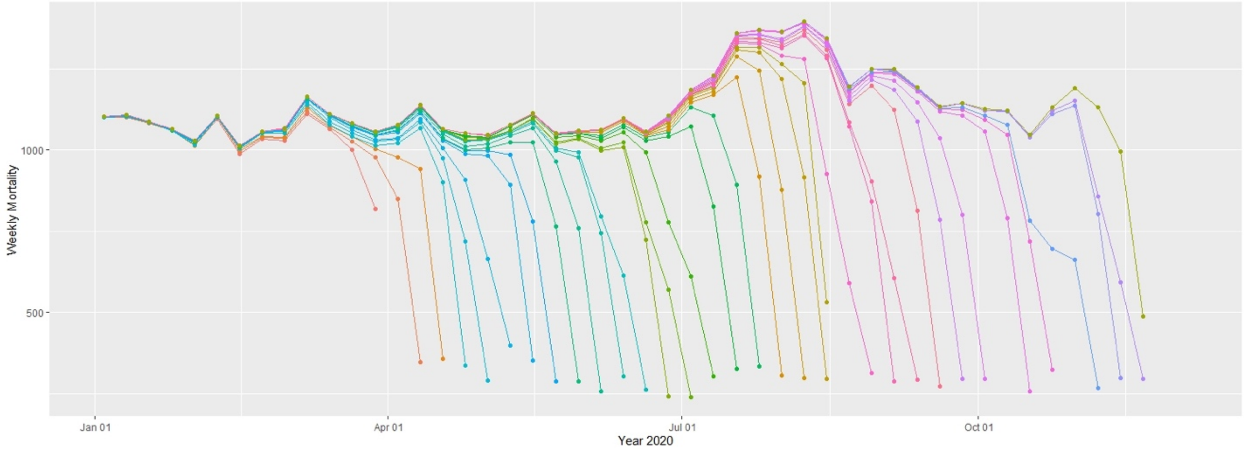

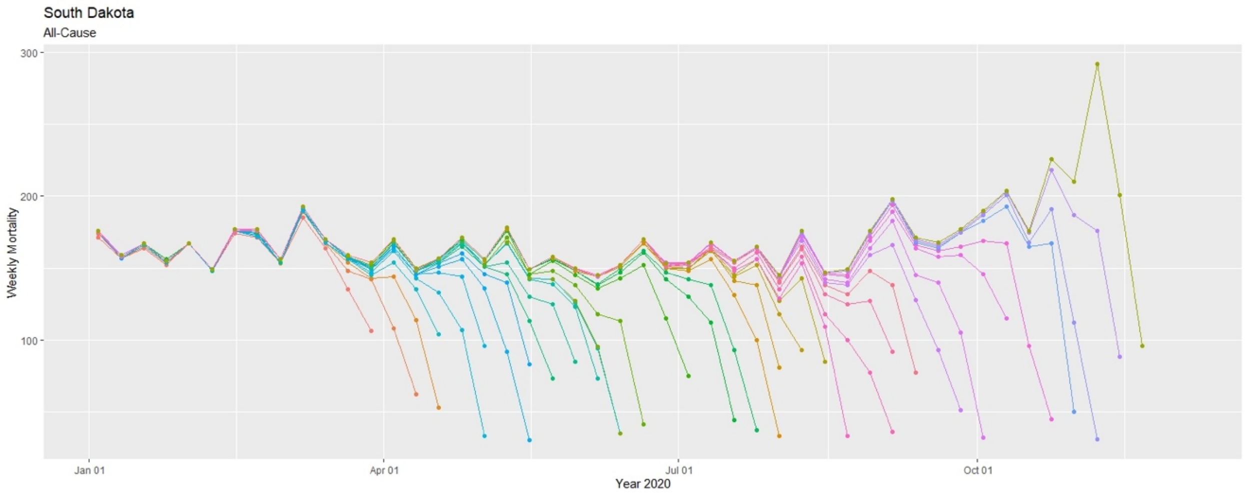
South Dakota
All-Cause
Weekly Mortality
300
200
100
Jan 01
Apr 01
Jul 01
Oct 01
Year 2020

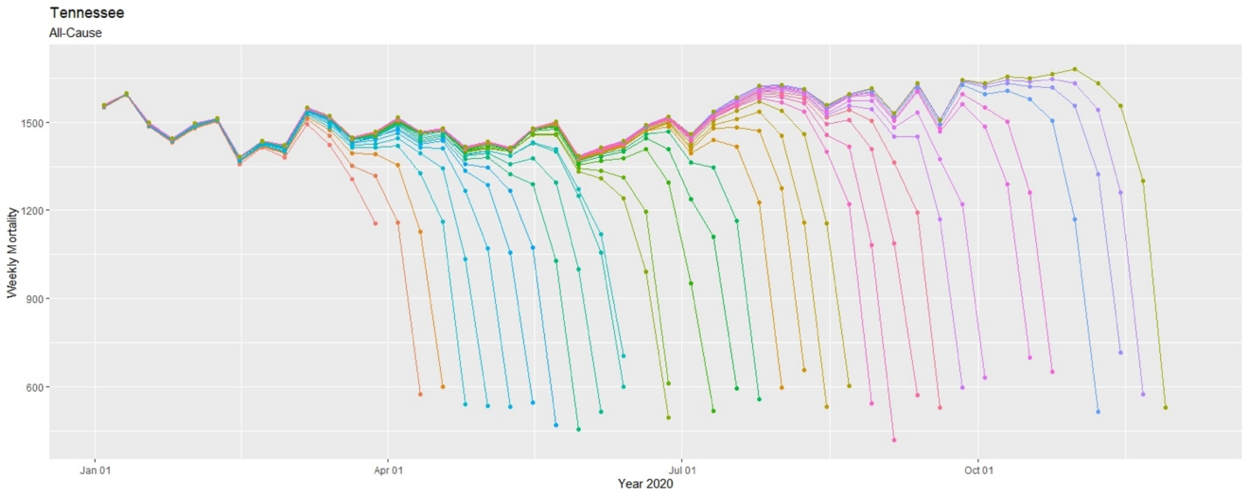
Tennessee
All-Cause
Weekly Mortality
1500
1200
900
600
Jan 01
Apr 01
Jul 01
Oct 01
Year 2020

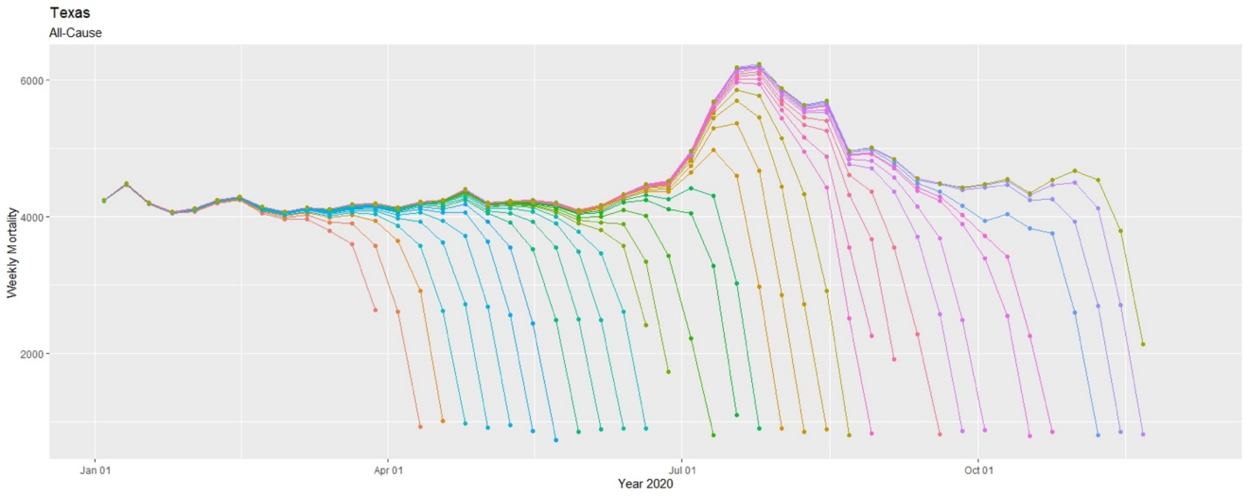
Texas
All-Cause
Weekly Mortality
6000
4000
2000
Jan 01
Apr 01
Jul 01
Oct 01
Year 2020

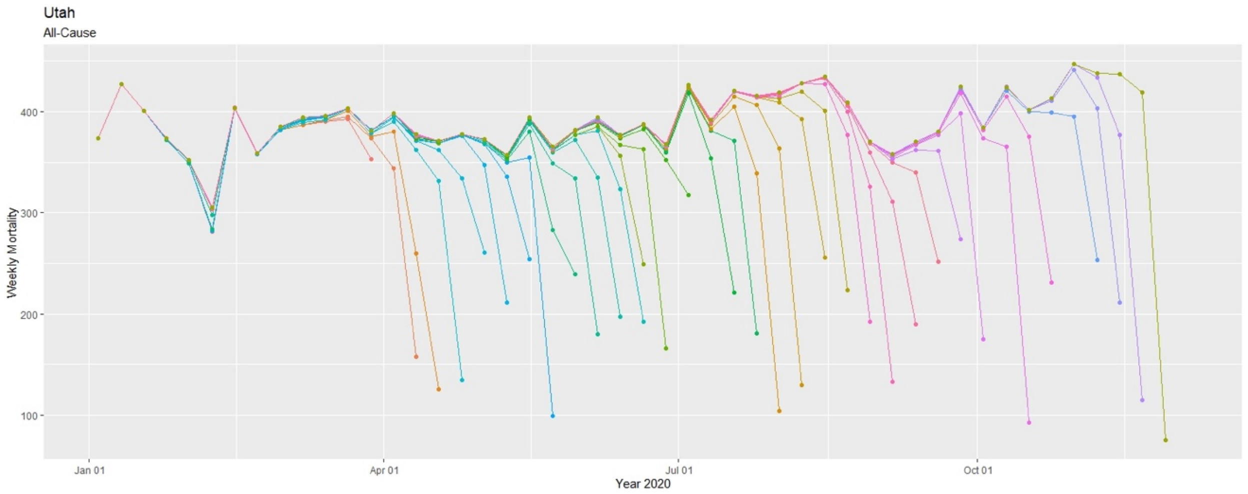
Utah
All-Cause
Weekly Mortality
400
300
200
100
Jan 01
Apr 01
Jul 01
Oct 01
Year 2020

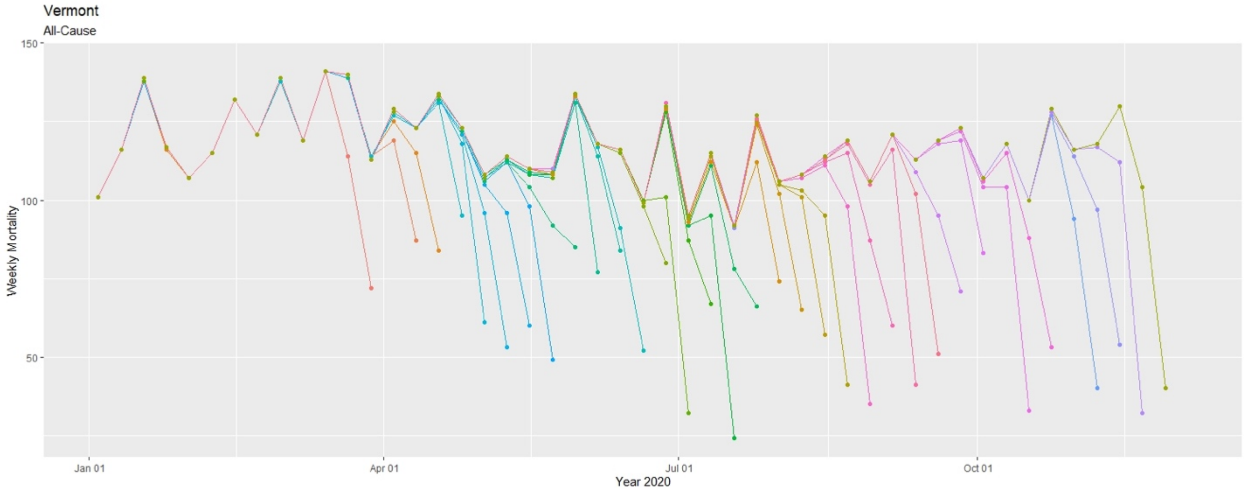
Vermont
All-Cause
Weekly Mortality
150
100
50
Jan 01
Apr 01
Jul 01
Oct 01
Year 2020

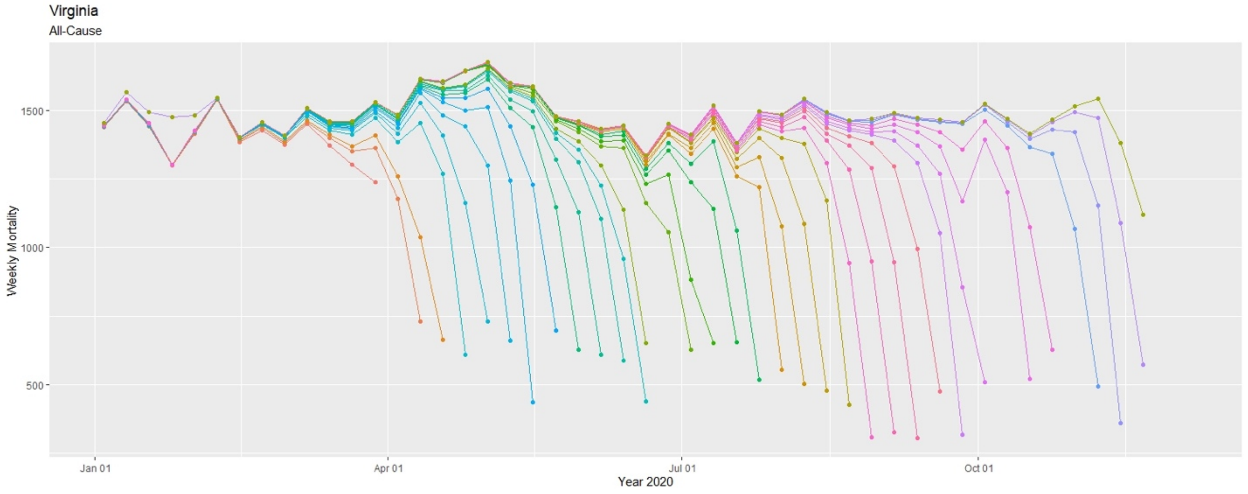
Virginia
All-Cause
Weekly Mortality
1500
1000
500
Jan 01
Apr 01
Jul 01
Oct 01
Year 2020

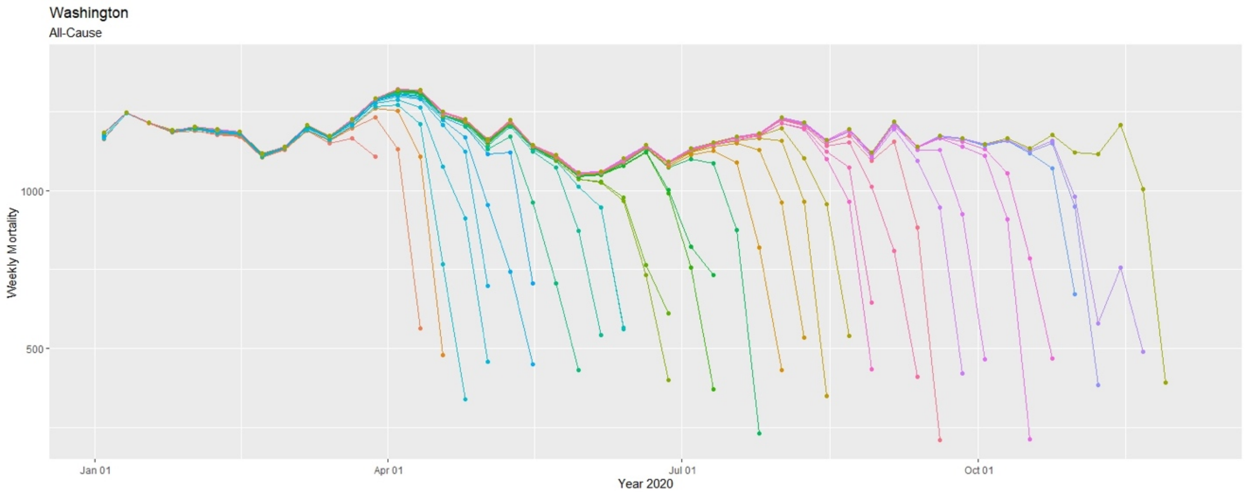
Washington
All-Cause
Weekly Mortality
1000
500
Jan 01
Apr 01
Jul 01
Oct 01
Year 2020

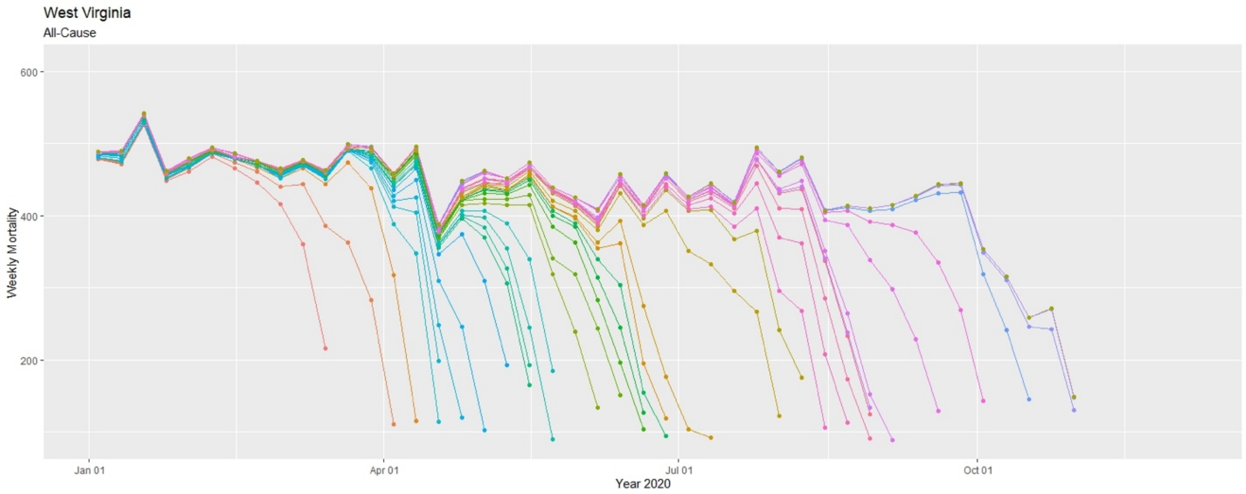
West Virginia
All-Cause
Weekly Mortality
600
400
200
Jan 01
Apr 01
Jul 01
Oct 01
Year 2020

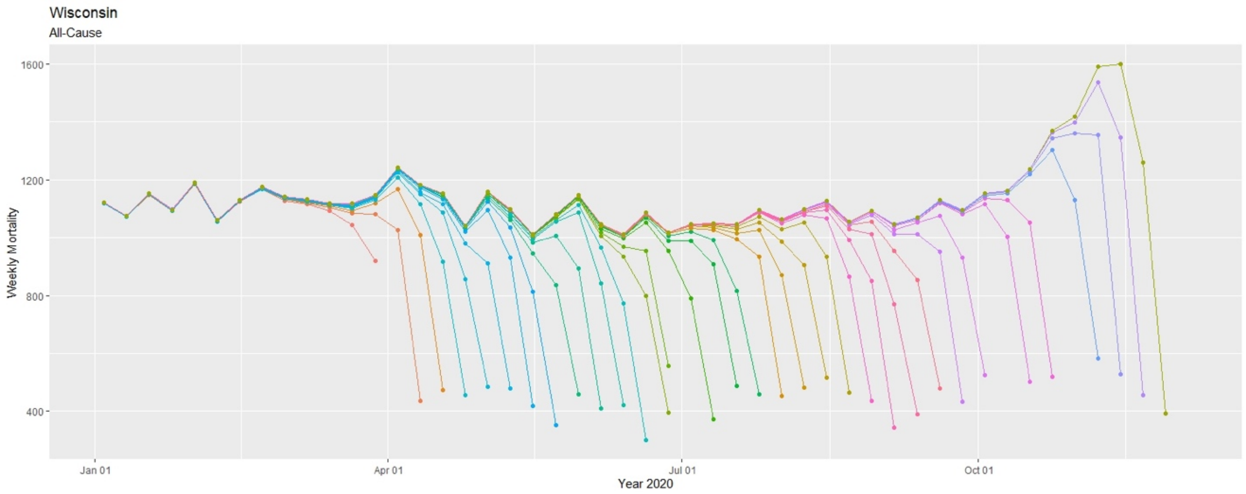

Wisconsin
All-Cause
Weekly Mortality
1600
1200
800
400
Jan 01
Apr 01
Jul 01
Oct 01
Year 2020


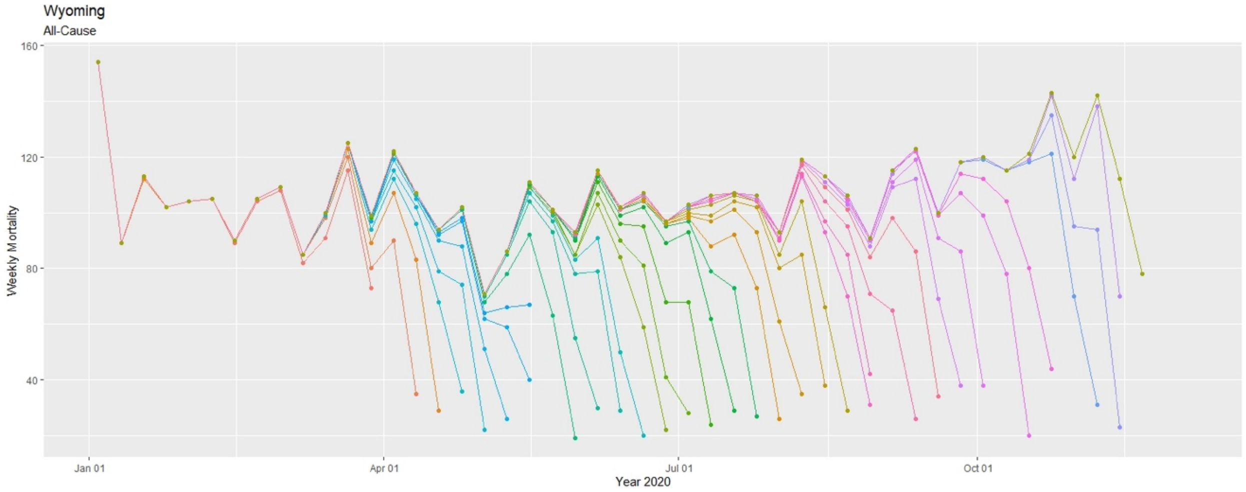

Wyoming
All-Cause
Weekly Mortality
160
120
80
40
Jan 01
Apr 01
Jul 01
Oct 01
Year 2020
